# Analysis of the Factors Affecting the Adoption and Compliance of the NHS COVID-19 Mobile Application: A National Online Questionnaire Survey in England

**DOI:** 10.1101/2021.03.04.21252924

**Authors:** Marcus Panchal, Sukhpreet Singh, Esther Rodriguez-Villegas

## Abstract

**Objectives:** To conduct an independent study investigating how adults perceive the usability, and functionality of the “NHS COVID-19” app. This study aims to highlight strengths, and provide recommendations to improve adoption of future contact tracing developments.

**Design:** A 60-item, anonymous online questionnaire, disseminated through social media outlets and email-lists by a team from Imperial College London.

**Setting:** England

**Participants:** Convenience sample of 1036 responses, from participants aged 18 and above, between December 2020 to February 2021.

**Primary Outcome Measures:** Evaluate the compliance and public attitude towards the “NHS COVID-19” app, regarding its functionality and features. This included whether participants expectations were met, and their thoughts on the app privacy and security. Furthermore, to distinguish how usability, perception, and adoption differed with varying demographics and user values.

**Results:** Fair app compliance was identified, with the app meeting expectations of 62.1% of participants who stated they downloaded it after weighted analysis. However, participants finding the interface challenging were less likely to read information in the app and had a lesser understanding of its functionality. Furthermore, lack of understanding regarding the app’s functionality and privacy concerns were possibly reasons why users did not download it. A readability analysis of the text revealed that app information was conveyed at a level which might only be accessible to under 60% of the population. The study highlighted issues related to the potential of false positives caused by the design choices in the “Check-In” feature.

**Conclusion:** This study showed that while the “NHS COVID-19” app was viewed positively, there remained issues regarding participants’ perceived knowledge of the app functionality, potentially affecting compliance. Therefore, we recommended improvements regarding the delivery and presentation of the app’s information, and highlighted the potential need for the ability to check out of venues to reduce the number of false positive contacts.

**Strengths and Limitations:**

- This is the first study assessing the perceived usability and functionality of the “NHS COVID-19” app directly from app users.
- Diverse sample size, with representation from all major regions of England.
- Statistical analysis to compare perceived app usability and functionality across demographics and the participants values regarding privacy and information.
- Study was conducted during lockdown where app use may have been minimal.
- Study may have missed potential participants who were not active on the relevant social media channels and email mailing lists which were used to disseminate the survey.

## Introduction

The Coronavirus (COVID-19), has become widespread, resulting in over 120,000 UK citizen deaths over the last 12 months [1]. Many unprecedented measures have been imposed on citizens day to day lives such as full isolation of all non-key workers, social distancing, and mask wearing. Contact tracing strategies, aiming at early identification and preventative quarantining of subjects who have been in close proximity of a confirmed virus carrier, have been up-taken by public health authorities to help further prevent the spread of the disease. It has been estimated that if 80% of contacts could be identified, stopping the progression of the pandemic would be extremely likely [2]. Contact tracing strategies have been attempted across the globe, with varying degrees of success [3, 4]. Although in an ideal scenario, one hundred percent of close contacts of one hundred percent confirmed cases would be promptly identified and socially isolated, in practical terms there are limitations to any method trying to achieve this. These limitations include: lack of professional human resources for both interviewing patients and following up with the identified contacts; reliance on patients ‘memories for up to a two week time span; significant time delays between diagnosis and isolation of contacts; and potential violation of privacy. Furthermore, the resources required for manual contact tracing are significant, with projected costs of the UK’s national contact tracing solution (Track and Trace), surpassing £10 Billion [5].

The Track and Trace system was released as the UK began to exit its first national lockdown, requiring citizens to manually check in to venues by providing their name and contact information [6]. Citizens could then be alerted if an outbreak was reported at a venue they recently entered. Logistically, this approach meant that the staff in the venue were responsible for holding citizens accountable for providing their information upon entering. Although practical, this approach was clearly privacy intrusive, as businesses were provided with personal information of all visiting individuals. This process was also seen as incomplete, as the Track and Trace method is ineffective in public areas (Public Transport, Parks, etc.) where citizens cannot check in, and will likely encounter significantly more contacts than they can remember. To this end, smartphone-based digital contact tracing presented itself as a faster and potentially more efficient method of contact tracing, especially at larger scales [7, 8].

Countries such as South Korea, Singapore, and Taiwan showed promise early on in the use of digital solutions to combat COVID-19 [9]. In South Korea, for example, the location of every subject is continuously monitored via their mobile phone, allowing the possibility of identifying who has been in contact with a newly diagnosed individual and for what period of time preceding that diagnosis [10]. The positive effects of this approach, in terms of modulation of the epidemic curve, are proven by the fact that South Korea is the country in which the rate of transmission, after reaching a critically large number of infections, was the lowest in the world early in the pandemic [11]. But, adopting a location based approach poses many ethical questions [12–14]. Whilst this may be effective and justified in stages of the pandemic in which health services are under risk of collapsing; many public concerns have been raised regarding these system’s, due to their lack of personal privacy. For example, South Korea’s digital contact tracing solution provides the government with access to GPS locations, and card transaction logs of each citizen [15]. This type of approach drew many concerns in the UK, which were clear from early surveys suggesting citizens would not endure a contact tracing solution that is privacy and location intrusive [16, 17]. Thus, to improve uptake of the app, the UK government, in common with many European governments, adopted different design approaches in the development of the “NHS COVID-19” app, focusing on providing UK citizens with a more privacy preserving digital contact tracing solution.

The “NHS COVID-19” app aims to be a one-stop system, allowing users to book a COVID-19 test, to self-report a positive COVID-19 result, and also alerts them if they encountered another app user who tested positive for COVID-19. This app uses Bluetooth technology paired with a QR-code based venue check in system to dictate close contacts between users. Furthermore, the app has additional features: informing of the latest COVID-19 restrictions, providing a risk level associated with the users ‘postal code, and a symptom checker. To combat controversy surrounding privacy, this app has utilized a decentralized approach, claiming to take as little information as possible, and only requiring the users postal code when downloading [18, 19].

Thus far, the “NHS COVID-19” application has received over 20 million downloads, with over 1.7 million contacts being alerted regarding COVID-19 exposure from over 800,000 positive test results entered through the app [20]. However, it is predicted that uptake by approximately 40 million people is required for this app to have a significant impact on the pandemic [21, 22]. Despite respectable initial uptake, and claims of implementing a privacy preserving design, the development of the app has faced scrutiny due to development pitfalls, privacy, and security concerns which may have affected citizens overall perceptions of digital contact tracing [22–24]. For instance, it was recently reported that despite over 100 million instances occurring using the Venue Check in feature, as little as 284 exposure alerts were sent to 276 venues, suggesting possible inefficiencies in the system [25]. Furthermore, studies investigating uptake factors in other nations have shown that access to contact tracing apps is one of the largest factors affecting their uptake and use within vulnerable populations [26]. The aforementioned factors therefore may have been potential reasons for the “NHS COVID-19” app not having a higher level of adoption, which is ultimately crucial to maximize the effectiveness of the app [2]. However, no formal study has been carried out to explore the factors affecting this, specifically in relation to the “NHS COVID-19” app.

Studies have emerged, since the early mentions of a UK contact tracing app, to investigate likeliness of the technologies acceptance, and to explore factors affecting necessary compliance. Early in the pandemic, a study surveyed participants from France, Germany, Italy, the United Kingdom, and the United States to assess the potential acceptance of an app of this nature [17]. Of all participants, 1055 were from the UK, with 74% suggesting they would probably or definitely download a national contact tracing app, with main reasons for not downloading the app including government surveillance concerns, phone security, or increased anxiety of having this app [17]. Besides the study suggesting older participants were more concerned regarding their phone security, and less about government surveillance, no relationship was found between participant’s reasons not to download the app and their age or gender. Furthermore, Technology Acceptance Models (TAM) have been used to identify potential factors influencing citizens’ decisions on whether they will download the app [27]. An extended TAM was used to find that it is likely citizens would download the app regardless of privacy concerns, and the main factor in this decision was determined to be perceived usefulness of the app [28]. A similar study used a Health Belief Model (HBM), and found that perceived benefits of contact tracing apps was the largest factor for users downloading the app, followed by self-efficacy and perceived barriers [29]. Additionally, a report early in the pandemic suggested public trust and confidence, and technical design choices influencing the privacy and security of the system, would contribute largely to app adoption [30]. However, it was later suggested that privacy concerns might not have influenced the citizen’s perception of the app as was originally expected, possibly due to the large UK support towards the NHS [31].

Despite the large number of studies previously mentioned that have looked into public perception and application uptake, an investigation into public attitudes and user experience with application features post-release has not yet taken place for the “NHS COVID-19” app. This is therefore the first independent study that investigates how the features of this app are being used by the public, in addition to assessing how citizens perceive the app, both in regards to its functions and usability. The findings of this study highlight the strengths and shortcomings of the app based on users ‘perceptions and opinions. After this introduction, the paper reads as follows:

1. A Methods section to further inform the reader on the functionality behind the NHS COVID-19 app. Then the methodology behind the survey design, and the main themes being investigated including Compliance, User Values, Information Content, Usability, and Understanding of the app. Next the survey deployment strategy, and survey response metrics were outlined. Lastly, statistical methods used for analysis were provided.
2. The Results section highlights the survey results based on the survey questions related to each theme. Results were tabulated in the result section itself, and in the supplementary material. Statistical analysis results from the Chi-Squared, Kruskal Wallis and Multivariate Regression analyses were computed.
3. A Discussion section highlights how these results are relatable to the UK population, providing potential reasons for fallbacks of the app designs and recommendations for future designs of these systems. We then conclude with an overview of the limitations of our study.

## Methods

### NHS COVID-19 Application Features and Design

The “NHS COVID-19” app (Figure 1) contains a variety of features designed to support contact tracing within the population, booking testing services, and giving users an indication of the risk level in the area they live in.

**Figure 1:**
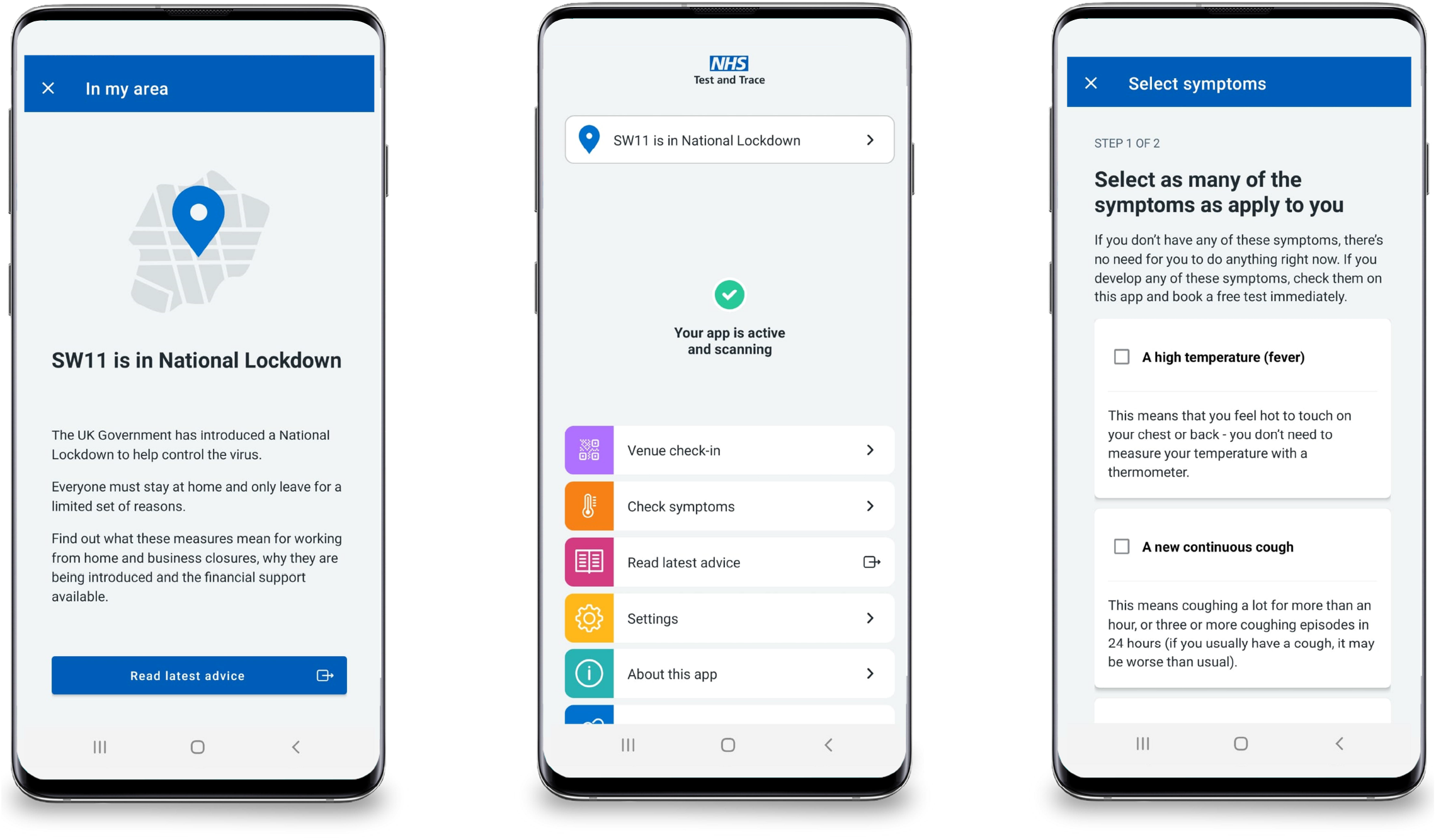
Sample screenshots of the “NHS COVID-19” app’s user interface [18,19].

Contact tracing is performed through two distinct services: an automatic proximity based system utilising the Bluetooth Low Energy Protocol (Bluetooth Contact Tracing); and a manual venue “check-in” system using the QR Code scanning features of mobile devices (Check-In). Automatic proximity based contact tracing is performed using an API developed jointly by Apple and Google, which requires each mobile device to constantly transmit a unique randomised identifying code (which is changed over a period of time) using the Bluetooth Low Energy Protocol. Each mobile device is also constantly scanning for nearby identifying codes from other devices from which distance can be estimated through the measure of Received Signal Strength Intensity (RSSI). This allows each mobile device to maintain an automatic log of other devices it has been within a certain distance from, which can then be used to inform users of those devices to self-isolate, if the user reports a positive test. Users have control on whether or not proximity contact tracing is enabled. The other method of contact tracing involves users manually scanning QR codes at venues they visit, to then store a log of the venues including the times visited. Users are checked out of venues when they either check into another venue or at midnight of the same day they checked in. Other users whose checked-in periods in the venue overlap in time will be notified of the need to self-isolate, if one of them reports a positive test.

The app also contains a symptom checker tool, designed to inform the users of what to do next depending on the symptoms they are experiencing, and allowing them to book a COVID-19 test. Finally, the app provides users with information on their COVID-19 infection risk within their general location, requiring the first half of the user’s postcode, as well as links to the latest COVID-19 advice.

### Survey Design

Our survey was designed to: 1) Determine how citizens perceived the usability of various features included within the “NHS COVID-19” app; 2) Ascertain the level of usage and compliance with these features; and 3) Evaluate the level of users’ understanding of the app.

To design our survey, we partly relied upon methodologies from various reviews and analyses of mobile health app usability studies, such as the Systems Wide Analysis of mobile health-related technologies (SWAT) [32]. The vast majority of these usability studies, however, apply a methodology across a wide range of health-related mobile apps, including questions which are either too general or refer to topics not relevant for this study. We therefore determined the common themes used to describe user perceived application usability within these studies, and generated focused questions specific to the “NHS COVID-19” application. The main themes used within our study are summarised in Table 1. To facilitate both the completion of the survey and also its analysis we used multiple choice questions (with users able to select a single option out of several), and where further detail was required, free text answer boxes. All questions were optional, and participants could go back, and revise answers throughout the survey, with questions not being randomized. Furthermore, we aimed to implement within our survey the recommendations from the CHERRIES checklist; a commonly used framework to design and implement online surveys in order to obtain high quality responses [33].

**Table 1:** Description of the main themes investigated in the study.

| **Study Themes** | **Description** |
| --- | --- |
| **Compliance** | Focused on how often users would use the previously outlined functions of the app within a given week or day, as well as identifying user habits that would facilitate the need to use these features. |
| **User Values** | Elicit the participant's feelings toward several issues that may influence their decision to use the app and its various features. These included participants own views on privacy as well as their beliefs on the privacy protection, security, and usefulness of the app itself. |
| **Information Content** | Sought to understand whether the "NHS COVID-19" application provided enough information to users about each feature within the app but also on the user's current status regarding their possible COVID-19 exposure, and the actions they should take based on this. |
| **Usability** | Encompassed the user's views on their ability to use the features of the app, whether these were simple or difficult to use, their ability to navigate the application and any technical difficulties they may have faced. |
| **Understanding the App** | Elicit participants' understanding of the basic functions and claims of the app. |

If participants did not download the “NHS COVID-19” app they were still able to complete the survey since this, on its own, helped to understand the reasons why they did not engage with the application to start with. Conditional formatting in the survey, however, ensured that participants’ usability based questions could not be answered by those who stated not to have downloaded the app. These participants were instead directed to a different set of questions that focused on trying to find out the reasons why they did not download it, and if any suggested changes would have changed their mind.

Design of the experiment was done by all contributing authors and the survey was pre-tested on a small cohort of institutional colleagues. Participants and members of the general public were not involved in the overall design, conduct, reporting, or distribution of the research plans.

### Survey Deployment

The study received ethical approval by the Science Engineering Technology Research Ethics Committee of Imperial College London (SETREC ref.: 20IC6427). The survey consisted of 60 questions which were implemented and released through the Qualtrics Xm online survey platform. The survey was open-access and spanned 18 pages, with 2-5 questions on each page. Qualtrics Xm ensured duplicate entries were not possible throughout the entirety of the survey through IP address checks. Before survey deployment, internal testing was done to iterate and improve the survey before release to the public. This survey was completely voluntary, and open to all members of the public in England who do not meet the exclusion criteria of being under 18 years of age. We obtained participants by non-probability (convenience) sampling through distributing our survey for 45 consecutive days (December 2020 and January 2021) via several different social media platforms (LinkedIn, Twitter, Reddit, Facebook, and NextDoor), official email lists to staff and students at Imperial College London, and word of mouth. We chose a non-probability sampling approach due to the low cost of obtaining responses from participants as well as the speed and ease at which participants could be recruited. Releasing our survey in a timely manner was an important factor due to the implementation of several lockdowns which severely affected the “NHS COVID-19” app’s use, requiring our survey to rely on participant recollection when providing responses. Therefore by releasing our survey in a timely manner, we were able to minimise any recall bias that may have occurred during data collection. Where surveys were deployed on social media these were either through official institutional accounts or the personal accounts of study organizers and shared by other accounts. Survey respondents needed to consent to take part in the study, and were required to be above the age of 18 as part of the inclusion criteria. Before providing consent, survey respondents were informed of the survey design, estimated time of completion, and data collection processes. Only completed surveys were used for analysis, other entries were deleted if not completed within 24 hours. The survey was intended to be distributed to as many subjects as possible throughout England and Wales and no incentives were offered to take part in the survey.

### Statistical Analysis

Due to our non-probability convenience sampling approach to our participant data collection, we are unable to accurately estimate the required sample size necessary in order to generalize our findings to the population of England and Wales. We therefore, aimed to receive at least 1000 respondents based on the scale of the sample size by similar studies [34–36].

The vast majority of our results were uni-variate with the proportion of responses for each category within these questions presented in the Results section below, and analyzed using descriptive statistics.

Additionally, we performed comparisons between questions, in order to identify significant correlations between certain user demographics/values/app-knowledge, with how users perceived the app and its features. Within these comparisons we performed the Chi-Squared test for independence on comparisons between categorical data and the Kruskal-Wallis H test between categorical and ordinal data with more than two groups. We evaluated the *p*-values for these comparisons and determined statistical significance at the level of *α*=0.05, performing Bonferroni correction where appropriate. In order to further quantify any relationships identified through statistical analysis we performed multivariate linear regression analysis. Categorical variables were encoded within our multivariate analyses using one hot encoding and age, gender, and geographical regions were used as fixed variables in all regression models. The analyses were carried out using the Python programming language with the commonly used Pandas, SciPy and Statsmodel Python packages.

### Textual Analysis

The Flesch Reading Ease test was used to assess the overall readability of the explanatory text in the on-boarding screens, and the “About” section of the app. Textual analysis was applied for each relevant screen, and included all explanatory text on the chosen screen. The Reading Ease test is based on the sentence and word lengths present in a body of text, disregarding word context, and is commonly used and accepted, both in research contexts and by regulatory bodies as part of the risk management process of manufacturers in medical devices certification [37–41].

## Results

### Participant Demographics

Out of 5967 participants who viewed the landing page, 1036 (17.4 %) of these participants who completed the study were used for analysis. The mean completion time was 10.0 minutes, with response times that were considered outliers (2 standard deviations less than the mean completion time) removed. The demographics of our sample can be seen in Table 2, and compared with the UK national demographics [42]

**Table 2:** Demographic and smartphone information of survey participants.

| Demographics |  |  |  |  |  |
| --- | --- | --- | --- | --- | --- |
| Question | Response(%) | ONS Statistics (%) | Question | Response (%) | ONS Statistic (%) |
| **Gender** |  |  | **Age** |  |  |
| Male | 589 (57.1%) | 21,662,879 (48.9%) | 18-25 | 245 (23.6%) | 5,487,272 (12.4%) |
| Female | 416 (40.3%) | 22,601,514 (51.1%) | 26-30 | 210 (20.4%) | 3,823,419 (8.6%) |
| Prefer Not to say | 27 (1.2%) |  | 31-40 | 275 (26.7%) | 7,500,184 (16.9%) |
| Other | 14 (1.3%) |  | 41-50 | 121 (11.6%) | 6,408,855 (14.5%) |
|  |  |  | 50+ | 185 (17.7%) | 21,043,663(47.5%) |
| **Region** |  |  | **Smartphone Usage** |  |  |
| London | 246 (23.6%) | 6,929,562 (15.7%) | 0-5 Years | 583 (56.5%) | 583 (56.5%) |
| South West | 105 (9.8%) | 7,210,838 (16.3%) | 5-10 Years | 440 (42.6%) | 440 (42.6%) |
| South East | 244 (23.5%) | 4,517,219 (10.2%) | 10+ Years | 9 (0.8%) | 9 (0.8%) |
| Midlands | 140(13.5%) | 13,357,128 (30.2%) |  |  |  |
| North East | 155 (14.9%) | 6,470,910 (14.6%) |  |  |  |
| North West | 121 (11.6%) | 5,777,736 (13.1%) |  |  |  |
| **Device OS** |  |  | **Education Level** |  |  |
| Android | 583 (56.5%) |  | Above A-levels | 711 (71.6 %) | 711 (71.6 %) |
| iPhone | 440 (42.6%) |  | A-level equivalent or below | 282 (28.3%) | 282 (28.3%) |
| Other | 9 (0.8%) |  |  |  |  |

Our analysis revealed differing proportions of demographics between our sample and the UK national demographics across age, gender, and location, likely due to self-selection biases, with the largest difference in the proportion of age 50+ participants sampled (17.7% vs. 47.5%). Therefore, we further processed our data to be more representative of all age groups, and regions in the UK by weighting the survey respondents, using a raking algorithm to generate the appropriate sample weights based on the true national demographic statistics for gender, age, and geographical region. We believe that by applying this weighting method, it allows us to more accurately generalize our findings to the UK population, and therefore all percentage results quoted in the body of the text in the following sections are the proportions after having weighting the samples unless otherwise stated. These methods are equivalent to a technique Lachan et al described to improve the quality of non-probability data of weighting the data set by raking [43].

Lastly, due to the multiple educational credentials submitted, education was grouped based on whether the participant’s highest educational achievement was above a UK A-level, or A-level and below.

### Compliance

A much larger proportion of participants (848, 84.1%) had downloaded the app, compared to those that did not (185, 15.9%). This proportion being significantly higher than the approximate 35% app uptake in England and Wales implies the study received more interest from participants who had downloaded the app. As per the reason 185 (15.9%) participants did not download the app, 68 (24.7%) of them “Did not feel safe downloading”, 67 (24.6%) “Did not see any benefit”, 29 (13.7%) stated “Due to place of work”, and 29 (20.2%) were unable to, due to the app not being compatible with their device, when asked the main reason they did not download the app. Furthermore, 74 (19.0%) of these participants responded that they would have downloaded the app “If it was completely anonymous”, despite the application already claiming to be completely anonymous.

When asked if participants changed their behaviour based on the assigned risk-level to their area, out of the users who downloaded the app, 519 (59.4%) marked that they had changed their behaviour, which indicated that the app had had some positive effect in terms of enabling users to make informed behavioural decisions, as a function of the local evolution of the pandemic.

The complete participant results outlining compliance can be found in Tables S1A and S1B.

### General App Usage

Among the 848 (84.1%) participants that did download the app, 722 (85.6%) of these participants were still using the app at the time of the survey, and 715 (81.6%) participants had the app downloaded for more than two months. The largest proportion of participants reported having opened the app once a week, 376 (42.6%). There was a significant proportion of users who had downloaded the app and did not engage with any of its foreground features, with 250 (25.2%) participants never opening the app after initially downloading, when asked how often they open the app. This may not be a concerning result however, as the app’s Bluetooth Proximity Contact tracing, one of the core features of the app, does not require regular app foreground use in order to function effectively. This does indicate though that a lot of the foreground functions may not be being used by a significant proportion of the population, such as the manual venue “Check-In” system, “Symptom tracker” and the “Read Latest Advice” tools. Lastly, results from the multivariate linear regression showed a positive correlation between users who are still using the app and feeling comfortable self-reporting a positive test (Table S14).

### Proximity Based Contact Tracing using BLE

Regarding the contact tracing features of the app, the majority of the participants (631, 76.5%) stated that they always kept the Bluetooth “Contact Tracing” feature enabled. While this is an encouragingly high proportion of users, this number not being 100% ultimately means that the number of app downloads may not be indicative of the amount of users who can be contact-traced via this method.

### Venue Check-in

When asked how many venues were visited on average per week, the majority of participants (447, 67.5%) stated they visited five or less venues a week before the November 2020 UK lockdown. However, of the participants able to visit venues, around half (494, 46.8%) indicated they used the “Check-In” feature between 75% to 25% of the time they visited a venue. Moreover, if participants forgot to check in, only 109 (16.5%) said they were reminded by a staff member, 450 (72.8%) indicated they were not reminded, and 67 (10.7%) used another method available to check-in. Thus despite participants more often than not using the “Check-In” feature at venues, these results indicate that venues could do more to engage with users to help fully utilise this feature, which may potentially also have the added benefit of increasing user venue check-in compliance. This can be further supported by the fact that 236 (40.0%) participants indicated that the NHS QR codes were accessible about half the time or less when visiting a venue.

When participants were asked how many venues on average they visited a day before the November 2020 UK lockdown, the majority said either zero (411, 52.8%) or one (311, 33.1%), with the rest indicating two or more (129, 14.0%). In addition to this, only 21 (1.6%) participants indicated that they would visit a venue for more than three hours on average. However, as the venue “Check-In” feature only checks users out when they visit another venue or at midnight on the same day, this could result in a large number of users being checked into venues much longer than they were actually present. This in turn may lead to a false number of encounters when cross-referencing between app users and venues they visited, thus possibly decreasing trust and confidence in the application’s ability to trace contacts accurately.

### Symptom Checker

Of the 224 participants who developed symptoms, many participants (151, 61.3%) entered these into the symptom checker tool within the app to obtain advice on how to proceed. This indicates that this feature was well used, thus contributing to more effective triage and management of users at a large scale.

### User Values

Participant results outlining user values, including their expectations of the app and views on privacy, can be found in Tables S2, S3.

### Privacy

Overall, 814 (75.9%) participants stated that they believed it is necessary for the app to protect the identities of individual users, with 531 (51.4%) participants noting that privacy in general is extremely important to them. Regarding the perceived privacy of the features, 589 (67.4%) and 613 (67.4%) participants were comfortable with the “Check-In” and the COVID-19 self-report features, respectively, in regards to privacy, and 531 (86.1%) respondents who downloaded the app noted they would self-report a positive COVID-19 test in the app if they were to test positive for COVID-19. This indicates that privacy is an important value for users, and the approach of the “NHS COVID-19” application, in promoting and utilising anonymous data may have led to the vast majority of users being content with using the features it provides.

Participants’ views on privacy, in general, were statistically analysed against various themes of the study. A statistical significance was shown between the importance of privacy to participants (of which 531 (51.4%) subjects in total indicated that privacy was extremely important to them) and whether they did or did not download the app (*p <* 0.0001). Of the participants that downloaded the app, 47.8% indicated privacy is extremely important to them. However this percentage increased for participants that did not download the app, with 68.4% indicating they consider privacy very important. This shows that despite the app not collecting any personal information, there may still be concerns affecting trust in the app.

In addition to users ‘views on privacy, participants also indicated that they strongly believed this app should not be legally enforced upon users, with 567 (64.5%) participants stating they do not think they should be legally obligated to download the app. This shows that users ‘value the autonomy to choose whether or not they engage with nationwide automated contact tracing programs.

Multivariate linear regression results for privacy importance can be found in Table S10. These results showed users views on privacy were statistically correlated with whether users had downloaded the app, required more information about the app from outside sources, and gender, with those that required more information and had not downloaded the app being more likely to value privacy.

### User Expectations

Participants’ responses regarding their expectations of the App and the resulting weighted analysis can be found in Tables S4A, S4B, and S4C.

Overall, over half (503, 62.1%) of the participants who had downloaded the app, marked that the app had met their expectations, with 340 (37.9%) stating otherwise. This was further analysed to compare the perceived adequacy of app feature information within the app and whether the application met the user’s expectations. We found a statistically significant difference between the expectations of those who found there was enough information on how close contacts were derived and those who did not (*p <* 0.0001). These data indicate that if the user found the information adequate then they would be more likely to say the app met their expectations (78.0%); compared with those who thought there was not enough information (57.8%). This can be further quantified from our Multivariate Regression analyses (Table S9) that found that Inadequate Information on close contacts was strongly negatively correlated with App Expectations (Coefficient: −0.28).

In regards to usability, relationships were drawn between user’s expectations and if the user felt the text was simple and easy to read (*p <* 0.001); if they felt the app was easy to navigate through (*p <* 0.001); and how they rated the overall style/interface of the app (*p <* 0.001). 67.8% and 71.5% of app users whose expectations were met, felt the app was simple and easy to read, and intuitive to navigate through respectively. Conversely, only 31.2% and 28.4% of users whose expectations were not met found the text on the app easy to read, and intuitive to navigate. Furthermore, multivariate regression analysis (Table S9) found a positive correlation between participants whose expectations were met, and those that found the app’s navigation intuitive (coefficient = 0.41). This potentially indicates that the design and interface of the app had a sizeable effect on the user experience.

### Information Content

Participant results including the participants views on the apps information content can be found in Tables S5, S6A, S6B.

With regards to the perceived information content of the app we identified whether users felt adequate information was presented for them to garner an understanding of the app’s major features. Overall a large proportion of participants believed the amount of informatory text presented to explain the features within the app was adequate for their needs, with 436 (52.1%) and 327 (39.6%) participants finding the information adequate to understand the “Check-In” and “Enter Test Result” features respectively.

Furthermore, many participants believed they were not provided adequate information regarding how the app dictated whether they had encountered another app user with COVID-19, with 320 (32.9%) feeling the information was inadequate and 296 (37.7%) feeling the information was neither inadequate or adequate. We also enquired as to whether participants ever required information from outside sources to better understand the app’s function and features; 204 (58.8%) participants responded that this was the case. The aforementioned points raise a possible concern, as a lack of information, and reliance on outside sources may increase the likelihood of misconceptions regarding the app and decrease user’s trust in the reliability and accuracy of its features.

There was also a statistically significant difference between the expectations of users who had obtained information from outside the app with those who had not (*p <* 0.0001). Participants who did not require external information were more likely to have found the app met their expectations (54.1%), compared with those who did have to use external sources of information (37.8%). Lastly, it was found through multivariate linear regression (Table S13) that finding the app text simple and easy to read was negatively correlated with requiring information regarding the app from outside sources (Coefficient: −0.24) which is to be expected.

### App Text Analysis

Many statistical relationships were drawn when grouping between users that did, and did not read all the information presented in the app. It appeared that a higher percentage of participants aged 50+, totalling 89 (64.9%) participants did not read all information presented in the app compared to the younger groups (*p <* 0.0001). Furthermore, respondents that did not read all the information, also did not find the text as easy to read (*p* <0.001), and the app as intuitive (*p <*0.005), compared to the participants that read all available information. Lastly, a higher percentage of participants (57.4%) had a better understanding of what their current status regarding their COVID-19 exposure was that read all the information compared to those that did not (*p* <0.005).

A relationship was drawn between users that both read all the information in the app, and are still using the app (*p <* 0.0001), with a slightly higher percentage of users that read all information continuing to use the app (57.7%) compared with users who did not read all the information. Ultimately, it appeared that participants that read all the information had increased trust in the app. However, for a smaller proportion of users it may not have been as straightforward and intuitive due to the interface/design of the app, and the complexity of the text.

In addition to whether or not the user read all of the information within the app, we found that the text within the app scored on average a value of 68.8 on the Flesch Readability Ease test. This level is associated with US Grade 8-9 students which in the UK is equivalent to a Reading Level 2 (GCSE A* - C), with this level only achieved by 57% of the UK General Public [44]. This leads to the possibility that users may have not engaged fully with the information within the app due to difficulty in understanding and interpretation.

### Usability

Participants’ views regarding the perceived usability, and their overall understanding of the app can be found in Table S7.

When participants who had downloaded the app were asked how intuitive the app navigation was, many were satisfied with the design aspects, as 591 (71.6%) stated the app’s navigation was intuitive. Furthermore, many participants felt the style/interface was consistent across the app most of the time (377, 53.9%), and the views were generally favourable on how participants viewed the style/interface, with 377(46.5%), 269 (32.4%), and 75 (8.30%) indicating they somewhat liked, neither liked nor disliked, or somewhat disliked the interface of the app respectively.

Regarding the status indicating if the participant had encountered another app user with COVID-19, 554 (66.8%) participants marked it as clear what their current status was. However the proportion of users who were not always aware of their current status was not negligible and therefore this may contribute to uncertainty regarding their exposure.

In regards to power usage, only 115 (13.5%) felt the app was significantly impacting the performance of their battery.

### Understanding of the App

Participants’ responses regarding their knowledge of the app functions and weighted analysis can be found in Tables S4A, S4B, S4C, and S8.

In total, only 400 (39.1%) participants believed the app does not require the user’s personal information to function. Furthermore, on the multi-choice question asking what technology is used to identify close contacts, 298 (13.9%) falsely chose GPS. 887 (56.3%) and 573 (25.9%) correctly chose Bluetooth and self-check in logs respectively, which in fact are both used for detection. 197 (22.5%) participants chose “Yes” when asked if they believed the app could track them. 681 (73.4%) participants correctly indicated that venues are not provided with information when using the “Check-In” feature. Hence, there was a significant misunderstanding regarding some of the key features of the app, despite explanations for each of these features in the app itself. This may reflect on the app’s ability to effectively present and relay information to the user.

### App Knowledge Analysis

Out of all participants, 242 (26.3%) chose all answers correctly, 226 (24.5%) answered three correctly, 168 (18.2%) answered two correctly, 188 (20.4%) answered one correctly, and 96 (10.4%) did not answer any correctly. Based on these groups, a relationship was shown between test scores, and Android vs iOS users; with a higher percentage (30.1% vs. 20.1%) of Android users scoring perfect, and a lower percentage of Android users scoring a zero (8.7% vs. 12.6%).

Out of participants that did not download the app, only 20.0% of these participants answered either three or all questions correctly, compared to users who did download the app; with 52.7% of users answering either three or all answers correctly (*p <* 0.001). This is likely due to users that downloaded the app simply being presented with more information compared to users that did not. However, this could also allude to the possibility that misconceptions within the population may be the reason for a reduced uptake of the app.

In regards to usability, comparisons were drawn between app knowledge, and the simplicity to read the text (*p <* 0.001). Out of participants that scored higher (3 or 4), 57.0% and 55.9% respectively, found the app’s text easy to read and intuitive to navigate through. This again, seems to hint to the possibility of the app design being unable to effectively convey information to a smaller subset of participants, which may have hindered their ability to understand the fundamentals of the app. Results from the multivariate linear regression (Table S11) showed high correlation between the app knowledge score and still using the app, meaning it is likely a higher proportion of those that still use the app have an increased understanding compared to those that don’t. Furthermore, the analysis also showed a positive correlation with feeling the app is not taking more information than necessary.

### Technical Issues

Out of all of the participants that downloaded the app, 286 (19.1%) noted that they had experienced technical issues with the app, with 208 of these participants further explaining their difficulties in detail. These free text responses, regarding what participants considered technical issues to them, were broadly classified as shown in Table 3. It can be seen that the most common issue experienced was false exposure notifications (98, Unweighted 47.1%) and loading notifications. Given how the notification system is a critical feature of the app with regards to informing users of potential contacts, failures of this system may have led to reduced confidence in the contact detection system.

**Table 3:** Description of the technical issues raised by participants, as well as the number of participant responses

| **Technical Issues** |  |  |
| --- | --- | --- |
| **Reported Issue** | **Response (%)** | **Explanation** |
| **False Notifications** | 98 (47.1%) | Users citing that there was no extra information in the app regarding the false exposure notification, leaving them in doubt. |
| **Notification stuck on loading** | 48 (23.1%) | Notification being sent to users that would not provide any information, but simply load, often for a few hours. |
| **Exposure checks not downloaded** | 18 (8.7%) | Users suggested that the app would not download the exposure checks routinely. |
| **App Crash** | 13 (6.3%) | The app either stayed permanently on the blue introductory screen, or closed randomly during use. |
| **Unable to check in using the QR code** | 12 (5.8%) | A few users suggesting the app was unable to scan the QR code of a venue. |
| **Cannot change postal code** | 11 (5.3%) | The inputted postal code location was unable to be changed. |

### Improvements

In total, 467 (Unweighted 45.1%) comments were submitted when asked if participants had any general suggestions on the app. 326 (Unweighted 69.8%) of these responses could be grouped together into eight different classification categories as seen in Table 4.

**Table 4:** Summary of participant recommended additions to the “NHS COVID-19” app.

| **Improvements** |  |  |
| --- | --- | --- |
| **Suggestion** | **Response(%)** | **Explanation** |
| **Requesting more information regarding privacy and contact tracing** | 70 (24.3%) | Users mentioned they would benefit with additional information in relation to how a close contact is calculated, and where and how the data is being stored. Other suggestions included a more concise privacy policy with bullet points, an FAQ in the app, an in-app introductory video, and metrics pertaining to daily vaccination and COVID case updates for their specific zone. |
| **More information regarding contacts encountered** | 56 (19.4%) | Many users lacked trust when being told they needed to self-isolate, as they argued they came in contact with a minimal amount of others, or they did not leave their house. Furthermore, users suggested the app provide information regarding the duration of the contact, and/or the venue it occurred in. Suggestions included having a counter that refreshes each day showing how many encounters the app has registered, and having in-app support with NHS staff members to provide them with more certainty regarding their exposure to COVID19. |
| **Improved User Interface** | 46 (12.5%) | Many users were unaware of their current status, which caused uncertainty regarding whether the app was working or not. Suggestions included incorporating an indicator in the home-screen that the app has yet to identify adequate COVID-19 exposure to the user, and to its best knowledge that they are safe. Users were also uncertain whether the app was, or was not running in the background, with some users indicating the dynamic scanning icon on the home screen led them to believe the app only scans when open. Suggestions included the app provide more reminders that the app will in fact work in the background for less tech-savvy users. |
| **Venue Check Out** | 28 (15.1%) | Users were concerned that they could not check out of venues, and simply suggested an option to check out with the app. |
| **Increased compliance rate** | 37 (15.1%) | Some users were adamant the app should be mandatory to ensure compliance is maximized. Suggestions included inputting NHS ID. |
| **Use the Google & Apple API** | 36 (12.5%) | Many users were unaware that the "NHS COVID-19" app uses the Google/Apple API, and suggested that the Google/Apple API be used to ensure privacy is preserved and to increase public trust. |
| **Improved Notification System** | 31 (10.8%) | Many users were confused and uncertain when sent a false exposure notification, indicating that when they tapped the notification, they were not provided with more information. It was determined these notifications were sent by the Google/Apple API, however, users suggested this be addressed in the app introduction, and/or provide the user with a log of their exposure notifications. |
| **Allow for location change for users that travel to different zones** | 22 (7.6%) | Users that work across various locations in the UK, or moved during the pandemic did not find changing their inputted postal code to be trivial, and re-downloaded the app. Suggestions include allowing users to change zones in the app. |

## Discussion

Overall, the results demonstrated that users valued their privacy, and strongly believed that their identities should remain anonymous. To this end, many participants saw value in the privacy preserving digital contact tracing solution, with most users finding it useful, and meeting their expectations. Fair, although not optimum, compliance was noted regarding many of the apps features such as the venue “Check-In”, and enabling Bluetooth contact tracing. However, in order to maximize the benefits of digital contact tracing to fight the pandemic, maximizing users uptake is crucial, and there were clear indicators that usability, information content, and misconceptions of the app may have been responsible for the lack of understanding, and overall compliance. It is pivotal for any level of concern to be addressed, as recently it has been found that a 1% increase in uptake of digital contact tracing apps could reduce the number of cases by between 0.8-2.3% [45], highlighting the importance of understanding user perceptions of contact tracing apps so that uptake inhibiting factors can be addressed. It appeared usability was adequate for a subset of participants, as many participants that read all the information on the app found the text easy to read, and navigation to be intuitive. However, there was an inverse relationship for those that did not read all the information. This may be related to the readability score of the text only being comprehensible for roughly 57% of the population. There was a similar link with participants that found the app more usable, as they had an improved understanding of the app. Ultimately, it may be expected that by improving the overall user journey, and more effective conveyance of information suited for the whole population, overall understanding and compliance with app features may be improved. It was also clear that users that did not download the app had a poor understanding of key app features, with many believing the app uses GPS, and that it requires personal information. Although this is not clearly linked to the usability of the app, it could be linked to prior development setbacks, which may have ultimately affected public trust regarding security and privacy, or to the lack of information present in external sources such as newspapers, social media, ads, etc. These concerns which are specific to the UK public have already been highlighted in recent literature [31,34,46]. Furthermore, these concerns have also been evident with contact tracing solutions in other countries, with similar studies conducted in Australia and Switzerland suggesting government policy, trust, and privacy concerns have led to a potential drop in compliance [36,47].

The possibility of falsely identifying contacts, with the venue “Check-In” feature was evident, as once a user checks into a venue, they are not checked out until they visit another venue, or at midnight that same day. Many participants highlighted that their typical time spent at venues was often less than an hour and the number of venues visited within an average day was close to one. Hence, this is potentially a sizeable problem. A possible recommendation to help reduce this potential pitfall is to employ a “Check-Out’’ feature, allowing the user and/or venue to dictate when the user has left the premise, resulting in a more accurate representation of contacts encountered by the user. Another aspect of the Venue “Check-In” feature that can be improved upon is the engagement of both the venue and the user in the process of checking in. The current system relies mostly on the user to initiate the check in process, which can ultimately lead to unconscious noncompliance. By designing a system that requires both parties (venue and user) to engage with the process, compliance could be driven by either party.

Lastly, it was identified from commented suggestions that false exposure notifications, and aspects of the user interface were causing uncertainty in terms of users exposure; and this played a role in users confidence in the tool if they were told to self-isolate. Although a false exposure notification bug issue was addressed in the past (well before launching this study), it was clear that the app had limited information regarding this. Going forward, many participants suggested the app should provide an increased amount of information through optional FAQs, an introductory video, and increased feedback.

### Study Limitations

One of the major limitations of our work is the use of the Non-Probability/Convenience sampling approach to recruit participants for our survey. While this technique was used due to its low cost and rapid recruitment abilities, it also introduces self-selection bias within our data that makes it difficult to generalise any findings we have to the wider UK Population. We attempt in this paper to correct for some of this through sample weighting based on true National Age, Gender and Regional Population Demographics, however we acknowledge that there will be other bias ‘contained within our data that we are unable to account for (e.g non-response bias).

As our work endeavoured to investigate the compliance and usage of the main features and tools contained within the app, we ideally required the survey to be active at the same time as users were able to use the app outside their home. However due to the rapidly changing nature of the COVID-19 pandemic, not long after the “NHS COVID-19” app was released, the UK was placed into a 2nd and 3rd lockdown period with restrictions on movement and outside activity. We therefore tried to mitigate the effect of this on the results of our study by specifically referring users to how they would have used the app in the period between the first and second lockdown in all compliance questions involving the venue “Check-In” feature, which understandably was only able to be used during this time period. This may have introduced some recall bias within the data collected.

Further to this we aimed to release our survey within a short timeframe, aiming to reduce the amount of recall bias present within our data. This meant that we were unable to pre-test our survey amongst an initial group of independent test participants in order to fully assess any problems with our survey, both with any clerical errors or unclear question phrasing. This testing was instead carried out by members of our research team, however we acknowledge that a group of neutral test participants would have been a more robust method of validating our survey design before release.

Our method for distribution of the survey through social media and email accounts may have unintentionally biased our data towards participants with a higher technological understanding and skill. This may then have affected how easy participants found interacting with the app compared with those who we were unable to reach through our distribution system.

Our study also specifically focused on the “NHS COVID-19” app rolled out in England. Hence, not all of our conclusions might be applicable to other contact tracing apps rolled out around the world, not just because of their potentially different designs but also due to the different demographic characteristics of their particular geography.

Due to the limitations of survey based research and statistical tests with non-probability sampled data, we were not able to assess causality for any of the relationships found within our data, and instead are limited to determining correlation between variables, which however, we believe is still a useful metric to quantify to enable improvements in the app to be highlighted.

We also noticed that, as our study was advertised as an “Usability and Compliance” study, we received a significantly higher proportion of responses from people who had downloaded the app compared with those who had not. While this did not impact the usability and compliance sections of our research, a greater proportion of respondents who had not downloaded the app would have allowed us to derive stronger conclusions regarding reasons why members of the public had not installed the application, as well as the general views and beliefs this section of society holds.

Lastly, there were sections of the population we were unable to reach due to our web-based sampling approach. This includes vulnerable populations who suffer an increased risk of infection. It has previously been shown through a similar perception-based study that this section of society has a significantly reduced access to digital contact tracing solutions [26], therefore if we had a more representative sample of this population we could have looked in more depth at ways to improve their uptake of the app.

### Impact

This work reports an independent study demonstrating how the public perceived and used different features of the “NHS COVID-19” app through analysis of an UK wide survey. The results of this study can be used to inform future contact tracing development efforts in order to optimize the adoption, compliance and overall efficacy of digital tools.

## Supporting information

Cherries Checklist

Survey

## Data Availability

Due to restrictions in the ethics document, data beyond what is presented in this publication cannot be distributed.

## Data Sharing

Due to restrictions in the ethics document, data beyond what is presented in this publication cannot be distributed. If you have any questions, please email the corresponding author.

## Acknowledgements

The authors would like to express their sincere gratitude to those that assisted with the deployment of the survey, including techUK, and /r/coronavirusuk.

## Authors ‘contributions

MP, and SS led data collection, and survey distribution. MP and SS analyzed the data. MP, SS and ERV wrote the manuscript. ERV led the work. The authors declare no competing interests.

