## Supplementary Tables for "Analysis of the Factors Affecting the Adoption and Compliance of the NHS COVID-19 Mobile Application: A National Online Questionnaire Survey in England"

### Supplementary Information

### Tables

| Compliance | | | |
|---|---|---|---|
| **Question** | **Response (UW%, W%)** | **Question** | **Response (UW%, W%)** |
| **Downloaded the app** | | **Still using the app** | |
| Yes | 848 (82.1%, 84.1%) | Yes | 722 (85.1%, 85.8%) |
| No | 185 (17.9%, 15.9%) | No | 126 (14.9%, 14.2%) |
| **Reasons the app was not downloaded** | | **Reasons that would have made them download** | |
| The app was not available for the device | 19 (8.9%, 20.6%) | If made by a University | 19 (14.6%, 8.6%) |
| Did not feel safe downloading | 68 (32.1%, 24.7%) | If made by a Not-for-Profit Organisation other than a University | 29 (13.6%, 22.1%) |
| Did not see any benefit | 67 (31.6%, 28.4%) | If it was made by a third-party organization | 2 (0.94%, 1.25%) |
| Due to place of work | 29 (13.7%, 6.09%) | If it was completely anonymous | 74 (34.7%, 19.0%) |
| Other | 29 (13.7%, 20.2%) | Other | 29 (11.5%, 49.1%) |
| **Length of Time App has been Installed** | | **Read Privacy Policy** | |
| Under 2 Week | 19 (2.24%, 2.63%) | Yes | 271 (31.9%, 41.2%) |
| 2-8 Weeks | 114 (13.4%, 15.77%) | No | 578 (68.1%, 58.8%) |
| Over 2 Months | 715 (84.3%, 81.59%) | | |
| **How often was the app opened** | | **Was the ”Contact tracing” feature always enabled** | |
| Multiple Times a day | 13 (1.5%, 3.27%) | Always | 631 (74.4%, 76.5%) |
| Once a day | 46 (5.4%, 7.09%) | More than 50% of the time | 88 (10.3%, 6.85%) |
| Once every few days | 163 (19.2%, 21.9%) | Less than 50% of the time | 59 (6.9%, 8.45%) |
| Once a week | 376 (44.3%, 42.6%) | Never | 52 (6.1%, 6.43%) |
| Never | 250 (29.4%, 25.2%) | Unsure of Feature | 18 (2.1%, 1.79%) |

Table S1A: Survey question and answers regarding overall compliance with the ”NHS COVID-19” app. UW% is the Unweighted Sample Percentages, W% is the Weighted Sample Percentages.

| Compliance | | | |
|---|---|---|---|
| **Question** | **Response (UW%, W%)** | **Question** | **Response (UW%, W%)** |
| **How often the ”read latest advice” feature was opened** | | **Technical issues experienced with the app** | |
| Daily | 3 (0.4%, 1.1%) | Yes | 286 (33.7%, 29.1%) |
| Once every few days | 50 (5.9%, 8.6%) | No | 562 (66.3%, 70.9%) |
| Once a week | 199 (23.5%, 28.8%) | | |
| Never | 509 (60.0%, 52.5%) | | |
| Once | 87 (10.2%, 9.0%) | | |
| **If symptoms were developed, was the ”Check symptoms” feature used** | | **If you forgot to check in to a venue using the app, were you reminded by a staff member?** | |
| Yes | 151 (67.4%, 61.3%) | Yes | 109 (17.4%, 16.5%) |
| No, I forgot to use this feature | 40 (17.8%, 22.7%) | No | 450 (71.9%, 83.6%) |
| No, I was not aware of this feature | 33 (14.7%, 15.9%) | | |
| **Prior to this current lockdown, approximately how many venues were visited daily** | | **Approximate time spent at each venue visited** | |
| 0 | 411 (48.3%, 52.8%) | Less than 30 minutes | 298 (35.0%, 39.7%) |
| 1 | 311 (36.6%, 33.1%) | 30 minutes - 1 hour | 341 (40.0%, 40.1%) |
| 2 | 90 (10.6%, 8.6%) | 1-3 hours | 171 (20.1%, 17.1%) |
| 3 | 19 (2.2%, 2.2%) | More than 3 hours | 21 (2.5%, 1.6%) |
| 4 or more | 20 (2.4%, 3.2%) | No venues visited | 20 (2.4%, 1.6%) |
| **Prior to this current lockdown, approximately what percentage of venues in a week was the ” Check-in” feature use** | | **Prior to this current lockdown, how many venues would be visited a week** | |
| 100% | 213 (30.1%, 26.9%) | Less Than 5 | 588 (68.9%, 67.5%) |
| 75% | 167 (23.6%, 18.8%) | 5-10 | 207 (24.1%, 28.1%) |
| 50% | 120 (16.9%, 11.7%) | 10-15 | 40 (4.6%, 3.76%) |
| 25% | 162 (22.9%, 16.3%) | More than 15 | 13 (1.5%, 0.62%) |
| 0% | 45 (6.3%, 26.3%) | | |
| **Changed behavior based on assigned risk-level to area** | | **Read the information on all the screens in the app** | |
| Yes | 519 (61.1%, 59.4%) | Yes | 460 (54.2%, 51.1%) |
| No | 330 (38.9%, 40.6%) | No | 389 (45.8%, 48.9%) |

Table S1B: Survey question and answers regarding overall compliance with the ”NHS COVID-19” app. UW% is the Unweighted Sample Percentages, W% is the Weighted Sample Percentages.

| User Values | | | |
|---|---|---|---|
| **Question** | **Response (UW%, W%)** | **Question** | **Response(UW%, W%)** |
| **Believe it is necessary for the app to protect identities of users** | | **Felt the app is taking more information than required** | |
| Yes | 814 (78.9%, 75.9%) | Yes | 88 (8.5%, 5.9%) |
| Undecided | 132 (12.8%, 14.1%) | Undecided | 307 (29.8%, 34.6%) |
| No | 86 (8.3%, 9.9%) | No | 637 (61.7%, 59.5%) |
| **Comfort self-reporting a COVID-19 test in regards to privacy and security** | | **Comfort using the "Checkin" Feature in regards to privacy and security** | |
| Comfortable | 613 (71.9%, 67.4%) | Comfortable | 589 (69.4%, 67.4%) |
| Undecided | 177 (20.7%, 20.4%) | Neither comfortable nor uncomfortable | 172 (20.3%, 20.4%) |
| Uncomfortable | 62 (7.2%, 12.3%) | Uncomfortable | 88 (10.4%, 12.3%) |
| **Reason app was downloaded** | | **Likelihood subjects would self-report a COVID-19 test in the app** | |
| Curiosity | 117 (13.8%, 15.3%) | Very Likely | 531 (82.9%, 86.1%) |
| To help others | 506 (59.7%, 61.2%) | Neither likely nor unlikely | 482 (6.5%, 4.1%) |
| Marketing | 23 (2.7%, 3.8%) | Unlikely | 109 (10.6%, 9.8%) |
| To follow government instructions | 48 (5.6%, 4.8%) | | |
| Friend and family advice | 26 (3.7%, 4.9%) | | |
| Instructed by a venue | 73 (8.61%,9.9%) | | |
| **Was the app found to be useful** | | **Did the App meet expectations** | |
| Yes | 287 (33.8%, 32.2%) | Yes | 503 (59.7%, 62.1%) |
| Undecided | 293 (34.6%, 39.4%) | No | 340 (40.3%, 37.9%) |
| No | 269 (31.6%, 28.4%) | | |
| **How much privacy was valued** | | **Believed data collected by the app was secure** | |
| Extremely Important | 531 (51.5%, 51.4%) | Yes | 411 (48.2%, 46.5%) |
| Moderately Important | 482 (46.7%, 47.6%) | Unsure | 323 (37.9%, 15.2%) |
| Not Important | 19 (1.8%, 1.0%) | No | 118 (13.9%, 38.2%) |
| **Feel citizens should be legally obligated to download the app** | | | |
| Yes | 276 (26.7%, 21.2%) | | |
| Undecided | 189 (18.3%, 14.3%) | | |
| No | 567 (54.9%, 64.5%) | | |

Table S2: Survey question and answers regarding user-values related to the "NHS COVID-19" app. UW% is the Unweighted Sample Percentages, W% is the Weighted Sample Percentages.

| Unweighted Responses | Weighted Responses | | | | | | |
|---|---|---|---|---|---|---|---|
| | Privacy Importance | | | | Education | | |
| Compliance | Very | Moderate | Not | p-value | Above | Below | p-value |
| **Downloaded the app** | | | | | | | |
| Yes | 486(47.8%) | 470(50.1%) | 11(2.0) | ***p** < 0.0001** | 671(63.3%) | 255(26.7%) | $p > 0.01$ |
| No | 134(68.4%) | 88(30.4%) | 1(1.0%) | | 147(69.0%) | 56(30.1%) | |
| **Read Privacy Policy** | | | | | | | |
| Yes | 147(56.2%) | 108(42.9%) | 2(0.7%) | ***p** < 0.0001** | 267(68.7%) | 103(31.2%) | $p > 0.01$ |
| No | 242(43.9%) | 299(53.4%) | 13(2.5) | | 404(72.2%) | 152(27.7%) | |
| **Changed behavior based on assigned risk-level to area** | | | | | | | |
| Yes | 310(48.6%) | 253(49.6%) | 6(1.7%) | $p > 0.01$ | 361(67.9%) | 184(32.1%) | ***p** < 0.001** |
| No | 177(46.7%) | 216(50.1%) | 5(3.0%) | | 310(79.4%) | 71(20.6%) | |
| User Values | | | | | | | |
| **Believe it is necessary for the app to protect identities of users** | | | | | | | |
| Yes | 526(57.5%) | 356(41.3%) | 5(1.1%) | ***p** < 0.0001** | 606(71.1%) | 235(28.8%) | $p > 0.01$ |
| Undecided | 56(28.7%) | 132(67.4%) | 4(3.7%) | | 129(57.0%) | 56(43.0%) | |
| No | 37(29.0%) | 69(65.1%) | 4(5.8%) | | 83(74.1%) | 21(25.8%) | |
| Information Needs | | | | | | | |
| **Required information about the app from outside sources** | | | | | | | |
| Yes | 194(49.0%) | 187(49.0%) | 4(1.8%) | $p > 0.01$ | 293(75.8%) | 85(24.1%) | ***p** < 0.01** |
| No | 293(46.9%) | 282(50.9%) | 7(2.1%) | | 377(67.3%) | 170(32.6%) | |

Table S3: Analysis between if participants privacy importance, and education, against various recorded answers from the survey. The Bonferroni adjusted score equated to 0.01, any $p$ value below this was statistically significant as indicated by the bold font and asterisk. Percentages were calculated after weighting the samples.

| | Weighted Responses | | | | | | | | |
|---|---|---|---|---|---|---|---|---|---|
| | Did app meet expectations | | | App knowledge score | | | | | |
| **Demographics** | **Yes** | **No** | **p-val** | **0** | **1** | **2** | **3** | **4** | **p-val** |
| **Device OS** | | | | | | | | | |
| Android | 360(59.6%) | 207(40.3%) | $p > 0.0038$ | 43(8.7) | 101(20.4%) | 89(18.0%) | 112(22.6%) | 149(30.1%) | $p > 0.0038$ |
| iPhone | 245(59.7%) | 153(40.2%) | | 47(12.6%) | 78(20.9%) | 72(19.3%) | 100(26.8%) | 76(20.3%) | |
| **Compliance** | | | | | | | | | |
| **Downloaded the app** | | | | | | | | | |
| Yes | - | - | - | 87(9.7%) | 158(17.7%) | 191(21.5%) | 249(28.0%) | 203(22.8%) | $\boldsymbol{p^* < 0.0001}$ |
| No | - | - | | 36(21.8%) | 42(25.4%) | 54(32.7%) | 12(7.3%) | 21(12.7%) | |
| **Technical issues experienced with the app** | | | | | | | | | |
| Yes | 117(43.6%) | 151(56.4%) | $\boldsymbol{p^* < 0.0001}$ | 68(10.8%) | 129(20.5%) | 119(18.9%) | 169(26.7%) | 143(22.8%) | $p > 0.0038$ |
| No | 209(29.9%) | 488(70.0%) | | 18(7.0%) | 28(14.8%) | 72(17.0%) | 79(30.7%) | 60(30.3%) | |
| **Still using the app** | | | | | | | | | |
| Yes | 580(65.7%) | 255(34.2%) | $\boldsymbol{p^* < 0.0001}$ | 61(6.5%) | 127(15.5%) | 162(16.9%) | 222(28.3%) | 189(32.6%) | $\boldsymbol{p^* < 0.0001}$ |
| No | 58(24.8%) | 104(75.2%) | | 25(7.0%) | 30(10.9%) | 29(28.0%) | 27(30.7%) | 13(23.3%) | |
| **Usability** | | | | | | | | | |
| **Overall was the text on the app simply and easy to read** | | | | | | | | | |
| Easy | 489(71.8%) | 192(28.1%) | $\boldsymbol{p^* < 0.0001}$ | 45(7.2%) | 98(15.6%) | 127(20.2%) | 188(29.9%) | 170(27.1%) | $\boldsymbol{p^* < 0.001}$ |
| Neither easy nor difficult | 71(37.8%) | 116(62.1%) | | 38(15.8%) | 59(24.6%) | 60(25.1%) | 53(22.1%) | 29(12.1%) | |
| Difficult | 4(20%) | 16(80 %) | | 3(17.6%) | 1(5.8%) | 7(41.8%) | 3(17.6%) | 3(17.6%) | |
| **Overall was the app intuitive and easy to navigate through** | | | | | | | | | |
| Easy | 404(71.3%) | 162(28.6%) | $\boldsymbol{p^* < 0.0001}$ | 30(4.7%) | 103(16.2%) | 146(23.2%) | 184(29.0%) | 171(26.9%) | $p > 0.0038$ |
| Neither easy nor difficult | 77(36.1%) | 134(63.8%) | | 53(23.3%) | 49(21.5%) | 40(17.5%) | 58(25.4%) | 28(12.2%) | |
| Difficult | 1(3.1%) | 28(96.6%) | | 4(18.2%) | 5(22.7%) | 4(18.2%) | 6(27.3%) | 3(13.6%) | |
| **Education** | | | | | | | | | |
| Above A-Level | 580(62.6%) | 346(37.6%) | $\boldsymbol{p^* < 0.0001}$ | 108(10.8%) | 189(18.9%) | 243(24.3%) | 248(24.8%) | 212(21.2%) | $p > 0.0038$ |
| Below A-Level | 1(3.1%) | 16(96.6%) | | 10(25.6%) | 9(23.0%) | 0(0.0%) | 8(20.5%) | 12(30.7%) | |

Table S4A: Analysis between if whether the app did meet expectations of the participant, and their overall app knowledge score, against various recorded answers from the survey. App knowledge score is classified as the number of the app knowledge questions participants correctly answered. The Bonferroni adjusted score equated to 0.0038, any $p$ value below this was statistically significant as indicated by the bold font and asterisk. Percentages were calculated after weighting the samples.

| | Weighted Responses | | | | | | | | |
|---|---|---|---|---|---|---|---|---|---|
| | Did app meet expectations | | | App knowledge score | | | | | |
| **Demographics** | **Yes** | **No** | **p-val** | **0** | **1** | **2** | **3** | **4** | **p-val** |
| **Opinion on the style/interface of the app** | | | | | | | | | |
| Like a great deal | 86(86.0%) | 14(14.0%) | $p$*< 0.0001 | 37(12.9%) | 48(16.7%) | 65(22.7%) | 45(15.7%) | 91(31.8%) | $p$*< 0.01 |
| Like somewhat | 253(69.5%) | 111(30.3%) | | 25(6.1%) | 64(15.5%) | 100(24.3%) | 109(26.5%) | 113(27.4%) | |
| Like nor dislike | 128(51.2%) | 122(48.8%) | | 50(34.7%) | 21(14.5%) | 10(6.9%) | 36(25%) | 27(18.7%) | |
| Dislike somewhat | 15(20.2%) | 58(79.7%) | | 17(13.6%) | 18(20.6%) | 27(31.0%) | 12(13.7%) | 13(14.9%) | |
| Dislike a great deal | 0(0.0%) | 19(100%) | | 1(2.2%) | 5(11.1%) | 33(73.3%) | 3(6.7%) | 3(6.7%) | |
| **Believe app is affecting performance of their device** | | | | | | | | | |
| Yes | 51(40.1%) | 76(59.8%) | $p$*< 0.001 | 23 (19.5%) | 40(33.3%) | 18 (15.2%) | 21 (17.8%) | 16 (13.5 %) | $p$*< 0.0001 |
| Undecided | 182 (66.9 %) | 90 (33.0%) | | 35(15.1%) | 61(26.2%) | 52 (22.5%) | 34 (14.6%) | 50 (21.5%) | |
| No | 372 (65.7%) | 194 (34.2%) | | 27 (5.1%) | 55(10.3%) | 121 (22.7%) | 192 (36.2%) | 136 (25.2%) | |

Table S4B: Continuation of Table S4A. Percentages were calculated after weighting the samples. App knowledge score is classified as the number of the app knowledge questions participants correctly answered.

| | Weighted Responses | | | | | | | | |
|---|---|---|---|---|---|---|---|---|---|
| | Did app meet expectations | | | App knowledge score | | | | | |
| Demographics | Yes | No | p-val | 0 | 1 | 2 | 3 | 4 | p-val |
| **Felt the app is taking more information than required?** | | | | | | | | | |
| Yes | 15 (36.5%) | 26 (63.5%) | $p^* < 0.0001$ | 19(30.6%) | 19 (30.6%) | 10(16.2%) | 9(14.5%) | 5(8.0%) | $p^* < 0.0001$ |
| Undecided | 155 (60.7%) | 100 (39.2%) | | 68(17.6%) | 102(29.5%) | 105(22.2%) | 53(18.4%) | 36(12.3%) | |
| No | 435 (65.1%) | 233 (34.8%) | | 36(18.6%) | 77(28.0%) | 133(28.8%) | 199 (14.5%) | 184(9.9%) | |
| **Comfort using the "Check-in" feature?** | | | | | | | | | |
| Comfortable | 426 (64.5%) | 234 (35.5%) | $p > 0.0038$ | 52(8.7%) | 89(14.9%) | 121(20.2%) | 173 (28.9%) | 162 (27.1%) | $p > 0.0038$ |
| Neither comfortable nor uncomfortable | 126 (41.3%) | 179(58.6%) | | 10 (5.5%) | 32(17.8%) | 54(30.2%) | 54(30.2%) | 29 (16.2%) | |
| Uncomfortable | 5 (45.5%) | 6(55.5%) | | 12 (12.6%) | 36(37.8%) | 15(15.7%) | 21(22.1%) | 11(11.5%) | |
| Information Needs | | | | | | | | | |
| **Was information adequate to understand on how close contacts are determined?** | | | | | | | | | |
| Yes, Information was adequate | 220 (78.0%) | 62 (21.9%) | $p^* < 0.0001$ | 21 (8.4%) | 49 (19.4%) | 41 (16.4%) | 78 (31.2%) | 61 (24.4%) | $p > 0.0038$ |
| Neither adequate nor inadequate | 246 (66.1%) | 126 (33.8%) | | 26 (8.9%) | 54 (18.6%) | 80 (27.5%) | 83 (28.6%) | 47 (16.2%) | |
| No, information was inadequate | 137 (57.8%) | 100 (42.1%) | | 39(11.4%) | 54 (15.8%) | 68 (20.0%) | 86 (25.2%) | 93 (27.3%) | |
| **Required information about the app from outside sources?** | | | $p^* < 0.0001$ | | | | | | $p > 0.0038$ |
| Yes | 192 (31.9%) | 409 (68.0%) | | 39 (10.7%) | 57(15.7%) | 69(19.0%) | 110 (30.3%) | 88 (24.4%) | |
| No | 188 (52.2%) | 172 (47.7%) | | 47(9.0%) | 100(19.2%) | 121 (23.2%) | 138 (26.5%) | 114 (21.9%) | |

Table 4C: Continuation of Table S4A and S4B. Percentages were calculated after weighting the samples. App knowledge score is classified as the number of the app knowledge questions participants correctly answered.

| Information Needs | | | |
|---|---|---|---|
| **Question** | **Response (UW%, W%)** | **Question** | **Response (UW%, W%)** |
| **Did the app provide adequate information to understand the ” Check-in” Feature?** | | **Did the app provide adequate information to understand on how to self-report a COVID19 test?** | |
| Yes, Information was adequate | 436 (51.4%, 52.1%) | Yes, Information was adequate | 327 (38.5%, 39.6%) |
| Neither adequate nor inadequate | 204 (24.0%, 21.6%) | Neither adequate nor inadequate | 90 (10.6%, 12.7%) |
| No, information was inadequate | 103 (12.1%, 11.9%) | No, information was inadequate | 64 (7.5%, 6.12%) |
| N/A, I did not use this feature | 106 (12.5%, 14.4%) | N/A, I did not use this feature | 368 (43.4%, 41.5%) |
| **Did the app provide adequate information to understand on how close contacts are determined?** | | **Required information from outside the app to improve understanding of app function?** | |
| Yes, Information was adequate | 232 (27.4%, 29.3%) | Yes | 376 (55.7%, 41.2%) |
| Neither adequate nor inadequate | 296 (34.9%, 37.7%) | No | 204 (44.3%, 58.8%) |
| No, information was inadequate | 320 (37.7%, 32.9%) | | |

Table S5: Survey question and answers regarding overall information content on the ”NHS COVID-19” app. UW% is the Unweighted Sample Percentages, W% is the Weighted Sample Percentages.

| | Weighted Responses | | | | | |
|---|---|---|---|---|---|---|
| | Read all the information in app | | | Required further information from outside sources | | |
| Demographics | Yes | No | p-Value | Yes | No | p-value |
| **Age** | | | | | | |
| 18-25 | 114 (55.6%) | 91 (44.4%) | | 110 (53.7%) | 95 (46.3%) | |
| 26-30 | 101 (58.4%) | 72 (41.6%) | | 87 (50.3%) | 86 (49.7%) | |
| 31-40 | 144 (61.8%) | 89 (38.2%) | **$p$*< 0.0001** | 103 (44.2%) | 130 (55.8%) | $p$> 0.005 |
| 41-50 | 53 (53.0%) | 47 (47.0%) | | 55 (55.0%) | 45 (45.0%) | |
| 50+ | 48 (35.0%) | 89 (64.9%) | | 45 (32.8%) | 92 (67.2%) | |
| Compliance | | | | | | |
| **Read Privacy Policy?** | | | | | | |
| **Yes** | 228(60.0%) | 152(40.0%) | **$p$*< 0.0001** | 229 (39.3%) | 358 (57.6%) | $p$> 0.005 |
| No | 251 (42.7%) | 336 (57.4%) | | 156 41.03%) | 224 (58.9%) | |
| **Still using the app?** | | | | | | |
| Yes | 393 (56.5%) | 297(43.0%) | **$p$*< 0.0001** | 329(39.3%) | 507(60.6%) | $p$> 0.005 |
| No | 41(34.1%) | 79(65.8%) | | 57(47.5%) | 63(52.5%) | |
| Information Needs | | | | | | |
| **Did the app provide adequate information to understand on how to self-report a COVID-19 test?** | | | | | | |
| Yes, Information was adequate | 204 (64.8%) | 113(35.2%) | | 136(43.1%) | 181 (56.9%) | |
| Neither adequate nor inadequate | 45 (56.2%) | 38 (43.8%) | **$p$*< 0.0001** | 25(41.6%) | 36(58.4%) | $p$> 0.005 |
| No, information was inadequate | 29(46.9%) | 32(53.1%) | | 50 (60.9%) | 33(39.1%) | |
| N/A, I did not use this feature | 156(45.7%) | 194 (54.3%) | | 150(42.9%) | 200 (57.1%) | |
| **Did the app provide adequate information to under-stand the "Check-in" Feature?** | | | | | | |
| Yes, Information was adequate | 262 (60.1%) | 174 (39.9%) | | 167 (38.3%) | 269 (61.7%) | |
| Neither adequate nor inadequate | 103 (50.5%) | 101 (49.5%) | **$p$*< 0.001** | 104 (51.0%) | 100 (49.0%) | **$p$*< 0.0001** |
| No, information was inadequate | 50 (48.5%) | 53 (51.5%) | | 59 (57.3%) | 44 (42.7%) | |
| N/A, I did not use this feature | 45 (42.9%) | 60 (57.1%) | | 45 (42.9%) | 60 (57.1%) | |
| **Did the app provide adequate information to understand on how close contacts are determined?** | | | | | | |
| Yes, Information was adequate | 149(67.4%) | 72 (32.5%) | **$p$ *< 0.0001** | 82 (37.1%) | 139 (62.8%) | **$p$ *< 0.0001** |
| Neither adequate nor inadequate | 125 (52.5%) | 113 (47.4%) | | 105 (36.7%) | 181(63.2%) | |
| No, information was inadequate | 160 (52.6%) | 144(47.6%) | | 168 (55.2%) | 136 (44.7%) | |

Table S6A: Analysis between participants that read all the information in the app, and participants information about the app from external sources, against various recorded answers from the survey. The Bonferroni adjusted equated to 0.005, any $p$ value below this was statistically significant as indicated by the bold font and asterisk. Percentages were calculated after weighting the samples.

| | Weighted Responses | | | | | |
|---|---|---|---|---|---|---|
| | Read all the information in App | | | Required further information from outside sources | | |
| Usability | Yes | No | P-value | Yes | No | p-value |
| **Overall, was the text simple and easy to read?** | | | | | | |
| Easy | 344(57.7%) | 257(42.2%) | $p* < 0.001$ | 236(39.3%) | 365(60.6%) | $p* < 0.0001$ |
| Neither easy nor difficult | 78(41.2%) | 111(58.7%) | | 103(54.4%) | 86(45.5%) | |
| Difficult | 12(57.1%) | 9(42.8%) | | 16(76.1%) | 5(23.8%) | |
| **Overall was the app intuitive and easy to navigate through?** | | | | | | |
| Easy | 363(53.2%) | 319(46.7%) | $p* < 0.001$ | 219(38.4%) | 348(61.5%) | $p* < 0.0001$ |
| Neither easy nor difficult | 104(40.1%) | 155(59.8%) | | 115(53.7%) | 99(46.3%) | |
| Difficult | 13(52.0%) | 12(48.0%) | | 21(70.0%) | 9(30.0%) | |
| **Opinion on the style/interface of the app?** | | | | | | |
| Like a great deal | 33(33.0%) | 67(67.0%) | | 37(37.0%) | 63(63.0%) | |
| Like somewhat | 204(55.8%) | 161(44.1%) | $p > 0.005$ | 155(42.4%) | 210(57.5%) | $p* < 0.005$ |
| Neither like nor dislike | 67(67.0%) | 33(33.0%) | | 110(43.4%) | 143(56.5%) | |
| Dislike somewhat | 36(48.6%) | 38(51.3%) | | 38(51.3%) | 36(48.6%) | |
| Dislike a great deal | 9(47.3%) | 10(52.6%) | | 15(78.9%) | 4(21.0%) | |
| **Was it clear what the current COVID-19 exposure status was?** | | | | | | |
| Most of the time | 318(57.4%) | 236(42.2%) | $p* < 0.005$ | 225(40.6%) | 329(59.3%) | $p < 0.005$ |
| About half of the time | 213(81.9%) | 47(18.1%) | | 57(53.2%) | 50(46.7%) | |
| Rarely | 70(66.0%) | 36(33.9%) | | 93(49.7%) | 94(50.2%) | |

Table S6B: Continuation of Table S6A. Percentages were calculated after weighting the samples.

| Usability | | | |
|---|---|---|---|
| **Question** | **Response (UW%, W%)** | **Question** | **Response (UW%, W%)** |
| **Generally, how accessible were the NHS QR codes at venues** | | **Overall, how would you rate the style/interface of the app** | |
| Always | 166 (20.0%, 20.3%) | Like a great deal | 106 (12.5%, 11.0%) |
| Most of the time | 333 (40.2%, 39.7%) | Like somewhat | 377 (44.4%, 46.5%) |
| About Half the time | 98 (11.8%, 10.2%) | Neither like nor dislike | 269 (31.7%, 32.4%) |
| Sometimes | 118 (14.2%, 13.2%) | Dislike somewhat | 75 (8.83%, 8.30%) |
| Never | 20 (2.4%, 3.2%) | Dislike a great deal | 22 (2.59%, 1.80%) |
| N/A- I haven't used the QR Codes | 94 (11.3%, 13.4%) | | |
| **Believe app is affecting performance of their device** | | **Overall was the text on the app simply and easy to read** | |
| Yes | 115 (13.6%, 13.5%) | Easy | 627 (73.8%, 70.9%) |
| Undecided | 220 (25.9%, 26.4%) | Neither easy nor difficult | 201 (23.7%, 27.3%) |
| No | 513 (60.5%, 60.1%) | Difficult | 21 (2.5%, 1.81%) |
| **Overall was the app intuitive and easy to navigate through?** | | **Was it clear what the current COVID-19 exposure status was?** | |
| Easy | 591 (69.6%, 71.6%) | Most of the time | 554 (65.2%, 66.8% ) |
| Neither easy nor difficult | 225 (26.5%, 25.9%) | About half of the time | 108 (12.7%, 10.7%) |
| Difficult | 33 (3.9%, 2.53%) | Rarely | 187 (22.0%, 22.4%) |
| **Was the design/appearance of the app consistent across all screens in the app** | | **Was booking a Covid-19 test through the app straight forward** | |
| Always | 325 (38.3%, 35.2%) | Yes, the app walked me through the process | 73 (76.8%, 80.6%) |
| Most of the time | 377 (52.7%, 53.9%) | No | 22 (23.2%, 19.4%) |
| About half the time | 269 (4.9%, 5.71%) | | |
| Sometimes | 75 (3.5%, 5.03%) | | |
| Never | 22 (0.5%, 0.19%) | | |

Table S7: Survey question and answers regarding usability of the "NHS COVID-19" app. UW% is the Unweighted Sample Percentages, W% is the Weighted Sample Percentages.

| App Knowledge | | | |
|---|---|---|---|
| **Question** | **Response (UW%, W%)** | **Question** | **Response (UW%, W%)** |
| **Believe the app requires personal information to function** | | **What technology do you believe is used to identify close contacts** | |
| Yes | 363 (34.2%, 39.6%) | GPS | 298 (15.9%, 13.9%) |
| Maybe | 179 (16.8%, 21.3%) | Bluetooth | 887 (47.2%, 76.3%) |
| No | 400 (37.7%, 39.1%) | Wifi-logs | 121 (6.4%, 3.9%) |
| | | Self-check in logs | 573 (30.5%, 5.9%) |
| **Believes app can track specific location with the app** | | **Believe the venues checked into using the app are provided with personal information** | |
| Yes | 197 (20.6%, 22.5%) | Yes | 98 (10.8%, 11.4%) |
| Maybe | 219 (23.0%, 20.0%) | Maybe | 131 (14.4%, 15.3%) |
| No | 537 (56.4%, 57.4%) | No | 681 (74.8%, 73.4%) |

Table S8: Survey question and answers regarding knowledge of the "NHS COVID-19" app. UW% is the Unweighted Sample Percentages, W% is the Weighted Sample Percentages.

| App Meets Expectations Multivariate Regression | | F-Statistic = 19.5<br>$r^2$ = 0.44 | | |
|---|---|---|---|---|
| **Question (Answer)** | **Coefficient** | **Std Error** | **T-Statistic** | **p-value (T-Statistic)** |
| Age (18-25) | 0.07 | 0.02 | 3.1 | 0.002 |
| Gender (Female) | 0.04 | 0.01 | 2.27 | .0006 |
| Read Latest Advice (Never) | 0.11 | 0.05 | 2.22 | .027 |
| Still Using app (Yes) | 0.19 | 0.05 | 4.0 | 0.0001 |
| Comfort with Self-Reporting Test (Neither comfortable nor uncomfortable) | 0.15 | 0.04 | 3.4 | 0.0001 |
| Navigation Intuitiveness (Easy) | 0.41 | 0.04 | 6.8 | 0.0001 |
| Navigation Intuitiveness (Neither easy nor difficult) | 0.10 | 0.04 | 2.3 | 0.02 |
| Navigation Intuitiveness (Difficult) | -0.19 | 0.05 | -3.6 | 0.0001 |
| Technical Issues (No) | 0.1412 | 0.045 | -2.8 | 0.006 |
| Read latest app advice (Never) | -0.14 | 0.05 | -.28 | 0.006 |
| Adequate information on close contacts (Inadequate) | -0.28 | 0.06 | -4.2 | 0.006 |
| Found text easy and simple to read (Easy) | 0.13 | 0.05 | 2.6 | 0.01 |

Table S9: Multivariate linear regression analysis investigating the effect of various variables in the prediction of whether the app met participants expectations. Variable coefficients and their associated p-values are presented along with the overall coefficient of determination for the linear regression model.

| Privacy Importance | F-statistic = 7.645<br>$r^2$ = 0.111 | | | |
|---|---|---|---|---|
| **Question (Answer)** | **Coefficient** | **Std Error** | **T-Statistic** | **p-value (T-Statistic)** |
| Age (26-30) | 0.162 | 0.05 | 3.25 | 0.001 |
| Gender (Female) | 0.2337 | 0.024 | 9.78 | 0.0001 |
| Gender (Male) | 0.2067 | 0.024 | 8.44 | 0.0001 |
| Downloaded the app (Yes) | 0.1477 | 0.014 | 10.4 | 0.0001 |
| Downloaded the app (No) | 0.2977 | 0.020 | 14.4 | 0.0001 |
| Read Privacy Policy (Yes) | 0.1477 | 0.014 | 10.4 | 0.0001 |
| Read Privacy Policy (No) | 0.2927 | 0.020 | 14.5 | 0.0001 |
| Changed behavior based on area risk level (Yes) | 0.0754 | 0.020 | 3.78 | 0.0001 |
| Required more information (Yes) | 0.3589 | 0.067 | 5.34 | 0.0001 |

Table S10: Multivariate linear regression analysis investigating the effect of various variables in the prediction of the importance of privacy for participants. Variable coefficients and their associated p-values are presented along with the overall coefficient of determination for the linear regression model.

| App Knowledge Multivariate Regression | F-Statistic = 14.45<br>$r^2 = 0.347$ | | | |
|---|---|---|---|---|
| **Question (Answer)** | **Coefficient** | **Std Error** | **T-statistic** | **p-value (T-Statistic)** |
| Downloaded the app (No) | 0.3258 | 0.069 | 4.732 | 0.0001 |
| Still using app (Yes) | 0.2297 | 0.047 | 4.856 | 0.0001 |
| Still using app (No) | -0.1234 | 0.051 | -2.429 | 0.015 |
| App met expectations (Yes) | 0.8128 | 0.287 | 2.836 | 0.005 |
| Information quality regarding self-reporting procedure (Inadequate) | -0.6515 | 0.200 | -3.265 | 0.001 |
| Accessibility of QR codes at venues (About half of the time) | 0.5836 | 0.186 | 3.130 | 0.002 |
| Feel app is taking more information than necessary (Yes) | 0.1969 | 0.098 | 2.000 | 0.046 |
| Feel app is taking more information than necessary (Undecided) | -0.2086 | 0.068 | -3.086 | 0.002 |
| Feel app is taking more information than necessary (No) | 0.4438 | 0.076 | 5.871 | 0.0001 |

Table S11: Multivariate linear regression analysis investigating the effect of various variables in the prediction of participants knowledge of the app. Knowledge of the app is classified as the number of the app knowledge questions participants correctly answered. Variable coefficients and their associated p-values are presented along with the overall coefficient of determination for the linear regression model.

| Read All Information Multivariate Regression | | F-Statistic = 6.678<br>$r^2 = 0.243$ | | |
|---|---|---|---|---|
| **Question (Answer)** | **Coefficient** | **Std Error** | **T-Statistic** | **p-value (T-Statistic)** |
| Still using app (Yes) | 0.15 | 0.05 | 3.0 | 0.0001 |
| Found text easy and simple to read (Difficult) | 0.23 | 0.067 | 3.58 | 0.001 |
| Navigation throughout the app (Difficult) | 0.11 | 0.035 | 3.364 | 0.001 |
| Read latest advice in app (Never) | -0.12 | 0.055 | -2.0 | 0.04 |
| Experienced Technical Difficulties (Yes) | 0.10 | 0.046 | 2.240 | 0.025 |
| Information regarding close contacts (Intuitive) | 0.10 | 0.04 | 2.8 | 0.004 |
| Felt app affected performance of their device (Yes) | -0.13 | 0.058 | -2.4 | 0.028 |

Table S12: Multivariate linear regression analysis investigating the effect of various variables in the prediction of whether the participants had read all the information within the app onboarding screens. Variable coefficients and their associated p-values are presented along with the overall coefficient of determination for the linear regression model.

| Required Extra Information Multivariate Regression | | F-Statistic = 8.452<br>$r^2 = 0.288$ | | |
|---|---|---|---|---|
| **Question (Answer)** | **Coefficient** | **Std Error** | **T-Statistic** | **p-value (T-Statistic)** |
| Still using app (Yes) | 0.23 | 0.052 | 4.593 | 0.0001 |
| Found text easy and simple to read (Easy) | -0.24 | 0.054 | -4.5 | 0.0001 |
| Found text easy and simple to read (Difficult) | 0.20 | 0.07 | 2.9 | 0.0001 |
| Experienced technical problems (No) | -0.19 | 0.046 | -4.2 | 0.0001 |
| QR Codes Accessible (Never) | -0.47 | 0.14 | -3.5 | 0.002 |
| Navigation throughout the app (Easy) | 0.15 | 0.042 | 2.9 | 0.004 |

Table S13: Multivariate linear regression analysis investigating the effect of various variables in the prediction of whether the participants required extra information from outside the app. Variable coefficients and their associated p-values are presented along with the overall coefficient of determination for the linear regression model.

| Still using app Multivariate Regression | F-Statistic = 43<br>$r^2 = 0.632$ | | | |
|---|---|---|---|---|
| **Question (Answer)** | **Coefficient** | **Std Error** | **T-Statistic** | **p-value (T-Statistic)** |
| Found text easy and simple to read (Neither easy nor difficult) | 0.17 | 0.03 | 6.8 | 0.0001 |
| App navigation (Intuitive) | 0.05 | 0.014 | .01 | 0.0001 |
| Believe app is taking more information than necessary (Yes) | 0.09 | 0.021 | 4.4 | 0.0001 |
| Required further information about the app (Yes) | 0.15 | 0.016 | 7.2 | 0.001 |
| Comfort with self reporting (Comfortable) | 0.24 | 0.02 | 10.2 | 0.002 |
| Comfort with self reporting (Not comfortable) | -0.12 | 0.03 | -3.6 | 0.004 |

Table S14: Multivariate linear regression analysis investigating the effect of various variables in the prediction of whether the participants were still using the app. Variable coefficients and their associated p-values are presented along with the overall coefficient of determination for the linear regression model.
