## Supplementary material for "Analysis of the Factors Affecting the Adoption and Compliance of the NHS COVID-19 Mobile Application: A National Online Questionnaire Survey in England": Survey

### Analysis of the Efficacy, Usability and User Perception of the NHS Covid-19 Mobile Application Version 1 – 11/11/20

---

Start of Block: Consent

Q60 **Participant Information sheet - Analysis of the Efficacy, Usability and User Perception of the NHS Covid-19 Application**

Thank you for considering to take part in this research study. Please read the following information to understand why this study is being conducted, and your involvement. If there is anything that is unsure, please contact one of the study investigators, contact details can be found in the bottom of this form.

**What is the main purpose of the study?**

There have been many initiatives throughout the country to help track and slow the spread of this devastating disease. Most recently, the NHS released the “NHS COVID-19” mobile application which aims to improve the efficiency of the prior Track and Trace system. This is done through a mixture of Bluetooth Proximity Sensing and QR Code Venue Scanning, allowing fast automated notifications to be sent out to users who need to self isolate.

Despite being downloaded by over 10 million individuals in the United Kingdom, an assessment of the effectiveness of the app, which is heavily dependent on user engagement, has yet to occur. Since this app is targeted towards the entire population, it is critical for this app to be intuitive, informative, and make users feel safe and secure while using the system.

This survey will be used to collect valuable insight and feedback on the “NHS COVID-19” mobile application. We will be asking you about your experience with the app, how often you use the app’s features, and your overall perception.

**Why have I been chosen?**

You were chosen to enter this study as we are looking for UK citizens over the age of 18 years old. You do not need to have downloaded and installed the NHS COVID-19 App in order to take part in this study.

**Do I have to take part?**

It is your decision whether you want to be involved with this study. By clicking “Accept” and continuing with the form, you are consenting to take part in this study. If at any time, you do not wish to proceed and would like to withdraw, click “cancel” or close the tab from your web browser.

If you take part, you simply have to complete the survey, that is all!

Once you submit your responses, you will be unable to withdraw the results, as all submissions are completely anonymous.

**What sort of questions will be asked?**

The survey is designed to further understand how users are experiencing different app features, if they were adequately informed within the app, how intuitive the app is, and how they generally perceive the app. Questions will be asked using a mixture of multiple choice options, check boxes, linear scales (e.g. Out of 1-5, how easy to use is this feature), and short free text boxes.

**Will my taking part in this study be kept confidential?**

To ensure your data’s protection and confidentiality a number of best-practice measures will be taken to ensure this is maintained. You will not be asked for any personal information. All your anonymous answers will be saved in a cloud utilizing the Google Forms platform, which can only be accessed through a user authentication system. Data will be held for 10 years post study end.

**What will happen to the results of the research study?**

If meaningful conclusions are obtained, the results are intended to be published in scientific journals.

**Who is organizing and funding the research?**

Imperial College London.

**What if I have complaints about this study?**

If you have any concerns about specific aspects of this study, in regards to the way you have been treated, then you should immediately contact Sukhi Singh – (Study co-investigator). If your response was not satisfactory, you may contact the Imperial College Research Governance Integrity Team.

**Who has reviewed the study?**

This study was reviewed by the Head of the Department and the Joint Research Compliance Office.

If you have any questions, please contact Marcus Panchal, otherwise, please continue.

Co-Investigator: Marcus Panchal

Department of Biomedical Engineering
South Kensington Campus
Imperial College London, SW7 2AZ

---

Q62 Please agree to all of the following statements below by ticking the corresponding boxes.

- ▢ 1) I am aged 18 Years or Older (1)
- ▢ 2) I confirm that I have read and understand the participant information sheet for this study and have had the opportunity to ask questions which have been answered fully. (2)
- ▢ 3) I understand that my participation is voluntary, and I am free to withdraw at any time, without giving any reason and without my legal rights being affected. (3)
- ▢ 4) I give consent for the information collection to be used to support other research in the future, including those outside of the European Economic Area (EEA) *Optional* (4)
- ▢ 5) I give permission for Imperial College to access my research records that are relevant to this research. (5)
- ▢ 6) I consent to take part in the above study (6)

*Skip To: End of Survey If Please agree to all of the following statements below by ticking the corresponding boxes. != 1) I am aged 18 Years or Older*

*Skip To: End of Survey If Please agree to all of the following statements below by ticking the corresponding boxes. != 2) I confirm that I have read and understand the participant information sheet for this study and have had the opportunity to ask questions which have been answered fully.*

*Skip To: End of Survey If Please agree to all of the following statements below by ticking the corresponding boxes. != 3) I understand that my participation is voluntary, and I am free to withdraw at any time, without giving any reason and without my legal rights being affected.*

*Skip To: End of Survey If Please agree to all of the following statements below by ticking the corresponding boxes. != 5) I give permission for Imperial College to access my research records that are relevant to this research.*

*Skip To: End of Survey If Please agree to all of the following statements below by ticking the corresponding boxes. != 6) I consent to take part in the above study*

**End of Block: Consent**

---

**Start of Block: Demographics**

Q1 What age group do you belong to?

- 18-20 (1)
- 21-25 (2)
- 26-30 (3)
- 31-40 (4)
- 41-50 (5)
- 50-65 (6)
- 65+ (7)

---

Q2 How long have you been using a smartphone for?

- 0 - 2 years (1)
- 3 -5 years (2)
- 6 - 8 years (3)
- 9 - 10 years (4)
- More than 10 years (5)

---

Q3 Are you currently using an iOS or Android device?

- Android (1)
- iOS (2)
- Other (3) ________________________________________________

---

Q4 What is your gender?

- ○ Male (1)
- ○ Female (2)
- ○ Prefer not to say (3)
- ○ Other (4) ________________________________________________

---

Q5 Which region did you spend the majority of your time during the last 30 days?

- ○ London (1)
- ○ South West England (2)
- ○ South East England (3)
- ○ West Midlands (4)
- ○ East Midlands (5)
- ○ Eastern England (6)
- ○ Yorkshire (7)
- ○ North West England (8)
- ○ North East England (9)
- ○ Other (10) ________________________________________________

**End of Block: Demographics**

---

**Start of Block: Background**

Q6 Do you believe it is necessary for the "NHS COVID-19" app to protect the identities of users?

- ○ Yes (1)
- ○ Maybe (2)
- ○ No (3)

---

Q7 Do you feel the "NHS COVID-19" app is taking more information than required?

- ○ Yes (1)
- ○ Maybe (2)
- ○ No (3)

---

Q8 Do you feel citizens should be legally obligated to download and use the "NHS COVID-19" app?

- ○ Yes (1)
- ○ Maybe (2)
- ○ No (3)

---

Q9 How important is your privacy to you?

- ○ Extremely important (1)
- ○ Moderately important (2)
- ○ Not at all important (3)

---

Q10 If you were to test positive for COVID-19, how likely would it be that you would self-report this in the app?

- ○ Very likely (1)
- ○ Neither likely nor unlikely (2)
- ○ Neither likely nor unlikely (3)

End of Block: Background

---

Start of Block: NHS App Use

Q11 Did you download the "NHS COVID-19" app?

- ○ Yes (1)
- ○ No (2)

---

*Display This Question:*

*If Did you download the "NHS COVID-19" app? = No*

Q12 If you did not download the "NHS COVID-19" app, was it for any of the below reasons? (Select all that apply):

- ▢ The app was not available for my device (1)
- ▢ I did not feel safe downloading this app (2)
- ▢ I believe this app will not benefit me (3)
- ▢ Other (4)

---

*Display This Question:*

*If Did you download the "NHS COVID-19" app? = No*

Q13 If applicable for you, would any of the below reasons changed your mind on downloading the app? (Select all that apply):

- ▢ If made by an University (1)
- ▢ If made by a Non-for-Profit Organisation other than a University (2)
- ▢ If it was made by a third party for profit (3)
- ▢ If it was completely anonymous (4)
- ▢ If it was downloadable on my device (5)
- ▢ If more information was provided about the app (6)

*Skip To: End of Block If Condition: If applicable for you, woul... Is Greater Than or Equal to 0. Skip To: End of Block.*

---

Q14 How long have you had the "NHS COVID-19" app downloaded on your device

- ○ Under 1 week (1)
- ○ 1-2 weeks (2)
- ○ 2-4 weeks (3)
- ○ 1-2 months (4)
- ○ Over 2 months (5)

---

Q15 Why did you first download the "NHS Covid-19" App?

- ○ I was curious (1)
- ○ I wanted to help others (2)
- ○ The marketing campaign convinced me (3)
- ○ I always follow what the government instructs me to do (4)
- ○ A friend or family member convinced me to (5)

---

Q16 Did the "NHS Covid-19" App meet your Expectations?

- ○ Yes (1)
- ○ No (2)

---

Q17 Do you find the "NHS Covid-19" App Useful?

- ○ Yes (1)
- ○ Maybe (2)
- ○ No (3)

---

Q18 Are you still using the "NHS Covid-19" App?

- ○ Yes (1)
- ○ No (2)

---

Q19 Did you ever delete the "NHS Covid-19" app for any reason?

○ Yes (1)

○ No (2)

---

*Display This Question:*

*If Did you ever delete the "NHS Covid-19" app for any reason? = Yes*

Q20 Why did you delete the "NHS Covid-19" app?

________________________________________________________________

________________________________________________________________

________________________________________________________________

________________________________________________________________

________________________________________________________________

---

Q21 How often do you open the "NHS Covid-19"?

○ More than once a day (1)

○ Once day (2)

○ Once every few days (3)

○ Once a week (4)

○ Never (5)

---

Q22 Do you always have the "Contact tracing" feature enabled?

- Always (1)
- More than 50% of the time (2)
- Less than 50% of the time (3)
- Never (4)
- I don't know what that is (5)

---

Q23 Prior to this current lockdown, how many venues (Restaurants, grocery stores, pubs, lectures halls, etc.) do you estimate you would visit in a given week?

- Less than 5 (1)
- 5-10 (2)
- 10-15 (3)
- More than 15 (4)

---

Q24 Prior to this current lockdown, approximately what percentage of these venues, in a given week, did you check into using the "Venue check-in" feature on the app?

- 100% (1)
- 75% (2)
- 50% (3)
- 25% (4)
- 0% (5)

---

*Display This Question:*

*If Prior to this current lockdown, approximately what percentage of these venues, in a given week, d... != 100%*

Q25 If at any point, you forgot to check into a venue using the "Venue check-in" app feature, were you immediately reminded by a staff member?

- ○ Yes (1)
- ○ No (2)

---

Q26 Prior to this current lockdown, on a typical day how many venues would you check into using the "Venue check-in" feature on the app?

- ○ 0 (1)
- ○ 1 (2)
- ○ 2 (3)
- ○ 3 (4)
- ○ 4 (5)
- ○ 5 or more (6)

---

Q27 Prior to this current lockdown, approximately how long would you spend at each venue you attended?

- ○ Less than 30 minutes (1)
- ○ 30 minutes - 1 hour (2)
- ○ 1-3 hours (3)
- ○ More than 3 hours (4)

---

Q28 Generally, how accessible were the NHS QR codes at venues you have attended?

- Always (1)
- Most of the time (2)
- About half the time (3)
- Sometimes (4)
- Never (5)

---

Q29 Do you believe this app is significantly impacting the performance, and/or reducing the battery life of your device compared to other apps you use?

- Yes (1)
- Maybe (2)
- No (3)

---

Q30 If you have developed any COVID-19 related symptoms since downloading the "NHS COVID-19" app, did you use the "Check symptoms" feature?

- Yes (1)
- No, I forgot to use this feature (2)
- No, I was not aware of this feature (3)
- No, I did not want to let anyone else know (4)
- No, I'd rather not say why (5)
- N/A - I have not developed symptoms since downloading the app (6)

---

Q31 How often did you open the “read latest advice” feature on the "NHS COVID-19" app?

- ○ Daily (1)
- ○ Once every few days (2)
- ○ Once a week (3)
- ○ Never (4)

---

Q32 Did you ever experience any technical issues/bugs with the app?

- ○ Yes (1)
- ○ No (2)

---

*Display This Question:*

*If Did you ever experience any technical issues/bugs with the app? = Yes*

Q33 What were these issues/bugs?

________________________________________________________________

________________________________________________________________

________________________________________________________________

________________________________________________________________

________________________________________________________________

**End of Block: NHS App Use**

---

**Start of Block: Usability**

Q34 Did the "NHS COVID-19" app provide adequate background information to fully understand the "Venue check-in" feature?

- ○ Yes, information was adequate (1)
- ○ Neither adequate nor inadequate (2)
- ○ No, information was inadequate (3)

---

Q35 Did the "NHS COVID-19" app provide adequate instruction on how to self-report a positive COVID-19 test directly from the app?

- ○ Yes, information was adequate (1)
- ○ Neither adequate nor inadequate (2)
- ○ No, information was inadequate (3)

---

Q36 Did the "NHS COVID-19" app provide adequate information on how the system determines if someone you encounter may be registered as a close contact?

- ○ Yes, information was adequate (1)
- ○ Neither adequate nor inadequate (2)
- ○ No, information was inadequate (3)

---

Q37 Overall, was the text on the "NHS COVID-19" app simple and easy to understand?

- ○ Easy (1)
- ○ Neither easy nor difficult (2)
- ○ Difficult (3)

---

Q38 Overall, was the "NHS COVID-19" app intuitive and easy to navigate through?

- Easy (1)
- Neither easy nor difficult (2)
- Difficult (3)

---

Q39 Overall, how would you rate the style/interface of the "NHS COVID-19" app?

- Like a great deal (1)
- Like somewhat (2)
- Neither like nor dislike (3)
- Dislike somewhat (4)
- Dislike a great deal (5)

---

Q40 Was the design/appearance of the "NHS COVID-19" app consistent across all the screens.

- Always (1)
- Most of the time (2)
- About half the time (3)
- Sometimes (4)
- Never (5)

---

Q41 Through the "NHS COVID-19" app, was it always clear to you what your current "status" was? E.g. Whether you have been in contact with someone who self-reported COVID-19?

- ○ Most of the time (1)
- ○ About half the time (2)
- ○ Rarely (3)

---

Q42 Were you told to self-isolate for 14 days?

- ○ Yes (13)
- ○ No (14)

---

*Display This Question:*

*If Were you told to self-isolate for 14 days? = Yes*

Q43 After receiving the notification that you needed to self-isolate, how clear were the instructions on what to do next?

- ○ Extremely clear (1)
- ○ Neither clear nor unclear (2)
- ○ Unclear (3)

---

*Display This Question:*

*If Were you told to self-isolate for 14 days? = Yes*

Q63 Did

- ○ Click to write Choice 1 (1)
- ○ Maybe (2)
- ○ No (3)

Q44 If you booked a COVID-19 test through the "NHS COVID-19" app, did you find this to be a straight forward process?

- ○ Yes, the app walked me through the process (1)
- ○ No, I was very confused (2)

---

Q45 Did you read all the information presented on the screens?

- ○ Yes, I read all the information (1)
- ○ No, I skipped through the screens (2)

---

Q46 Did you ever require information from outside the "NHS COVID-19" app (NHS website, news articles, social media, etc.) to further understand the app?

- ○ Yes (1)
- ○ No (2)

---

Q47 Do you understand how the "NHS COVID-19" app assigns "risk levels" to your area?

- ○ Yes (1)
- ○ No (2)

---

Q48 Did you ever change your behavior, based on the assigned risk-level to your area?

- ○ Yes (1)
- ○ No (2)

End of Block: Usability

Start of Block: App Security and Privacy

Q49 How comfortable in terms of privacy and security, were you with checking into venues using the "NHS COVID-19" app?

- Comfortable (1)
- Neither comfortable nor uncomfortable (2)
- Uncomfortable (3)

Q50 Do you believe data collected from the "NHS COVID-19" app is secure?

- Yes (1)
- Unsure (2)
- No (3)

Q51 How comfortable in terms of privacy and security are you with the self-reporting process in the "NHS COVID-19" app?

- Comfortable (1)
- Neither comfortable nor uncomfortable (2)
- Uncomfortable (3)

Q52 Did you read the privacy policy?

- Yes (1)
- No (2)

End of Block: App Security and Privacy

Start of Block: NHS App Function

Q53 I believe the "NHS COVID-19" app requires personal information to function. Please answer to the best of your knowledge.

- ○ Yes (1)
- ○ Maybe (2)
- ○ No (3)

---

Q54 What technology does the "NHS COVID-19" app use to register close contacts? Please answer to the best of your knowledge.

- ▢ GPS (1)
- ▢ Bluetooth (2)
- ▢ Wifi-Logs (3)
- ▢ Self-check in logs (4)

---

Q55 I believe the government is able to track my location using this app? Please answer to the best of your knowledge.

- ○ Yes (1)
- ○ Maybe (2)
- ○ No (3)

---

Q56 Venues I check into using the "NHS COVID-19" app are provided with my personal information? Please answer to the best of your knowledge.

- ◯ Yes (1)
- ◯ Maybe (2)
- ◯ No (3)

---

Q57 I believe that the "NHS COVID-19" app will alert me if someone I encountered, who was registered as a "close contact", self-reported a positive COVID-19 test. Please answer to the best of your knowledge.

- ◯ Definitely yes (1)
- ◯ Probably yes (2)
- ◯ Might or might not (3)
- ◯ Probably not (4)
- ◯ Definitely not (5)

**End of Block: NHS App Function**

---

**Start of Block: Improvements**

Q58 What would you have done differently in the "NHS COVID-19" app.

________________________________________________________________

________________________________________________________________

________________________________________________________________

________________________________________________________________

________________________________________________________________

---

Q59 Any further comments?

________________________________________________________________

________________________________________________________________

________________________________________________________________

________________________________________________________________

________________________________________________________________

**End of Block: Improvements**
